# Evaluation of stochastic trajectory-based epidemic models using the energy score

**DOI:** 10.1101/2025.01.13.25320493

**Authors:** Clara Bay, Kunpeng Mu, Guillaume St-Onge, Matteo Chinazzi, Jessica T. Davis, Alessandro Vespignani

## Abstract

Scoring rules are critical for evaluating the predictive performance of epidemic models by quantifying how well their projections and forecasts align with observed data. In this study, we introduce the energy score as a performance metric for stochastic trajectory-based epidemic models. As a multivariate extension of the continuous ranked probability score (CRPS), the energy score provides a single, unified measure for time-series predictions. It evaluates both calibration and sharpness by considering the distances between individual trajectories and observed data, as well as the inter-trajectory variability. We provide an overview of how the energy score can be applied to assess both scenario projections and forecasts in this format, with a particular focus on a detailed analysis of the Scenario Modeling Hub results for the 2023-2024 influenza season. By comparing the energy score to the widely used weighted interval score (WIS), we demonstrate its utility as a tool for evaluating epidemic models, especially in scenarios requiring integration of predictions across multiple target outcomes into a single, interpretable metric.

## Introduction

Epidemic models often generate probabilistic predictions, representing a range of plausible future trajectories rather than a single outcome. In contrast to deterministic forecasts based on fixed initial conditions, these probabilistic outputs incorporate uncertainty arising from random processes and variable parameter values (Sherratt et al., 2024). Clear communication of such uncertainty is essential for informing decision-makers about the spectrum of possible epidemic scenarios (McCabe et al., 2021).

Epidemic forecasting and scenario modeling initiatives, such as the CDC FluSight Forecasting Challenge, the COVID-19 Forecast Hub, and the Scenario Modeling Hub (SMH), require participating teams to submit probabilistic predictions in a quantile format.(Mathis et al., 2024; Cramer et al., 2022a; Howerton et al., 2023). Scoring rules, such as the weighted interval score (WIS), can then be applied to quantile format outputs to analyze the performance of projections with respect to observed surveillance data. Epidemic predictions, however, are often produced as collections of stochastic trajectories, each representing one possible realization of how an outbreak could evolve(Sherratt et al., 2024). Increasingly, collaborative forecasting hubs have requested these individual trajectories—rather than only aggregated quantile summaries—to provide greater transparency and a clearer view of the predicted epidemic dynamics. This shift, exemplified in recent rounds of the Scenario Modeling Hub, helps avoid the opacity of prediction intervals, which can mask important temporal features of the modeled outcomes(Loo et al., 2024).

The information contained within epidemic projections in a quantile format versus trajectory format has both advantages and disadvantages. Trajectories allow for greater flexibility in weighting and analysis of time-series features, where the quantile format does not retain information on variability within potential epidemic outcomes (Sherratt et al., 2024). Trajectories acknowledge the individual behavior of multiple potential realizations of an outbreak, where the quantiles evaluate the descriptive statistics at each time point. Fixed-time descriptive statistics can suppress important epidemiological information and obscure the true uncertainty of different epidemic outcomes (Juul et al., 2021; McCabe et al., 2021; Sherratt et al., 2024). Typically, predictions in a quantile format highlight the most likely outcomes, rather than the worst-case scenario, which is critically important to public health decision-makers (Wilke and Bergstrom, 2020). While this has implications for the communication of epidemic projections, it can also impact what we can learn from model evaluation.

Although the value of trajectory-based outputs is increasingly recognized, most evaluation methods for epidemic projections are still designed for quantile-based formats and have not been widely adapted to leverage the richer information trajectories provide. Here, we assess the utility of the energy score as a performance metric specifically tailored to evaluate epidemic model projections reported in stochastic trajectory format. The energy score has been applied across various fields, including weather forecasting (Gneiting et al., 2008; Kapoor et al., 2023), electricity market pricing (Cramer et al., 2023; Grothe et al., 2023), and wind/solar power generation (Pinson and Girard, 2012; Staid et al., 2017; Zhang et al., 2014; van der Meer, 2021). In epidemic modeling, the energy score has been applied in a limited number of cases to analyze multivariate time-series models (Held et al., 2017; Engebretsen et al., 2023; Bonacina et al., 2024). Moreover, computational packages used to assess probabilistic forecasts with proper scores have implemented the energy score (Jordan et al., 2019; **?**; **?**). However, it has yet to gain widespread adoption as a standard metric for evaluating epidemic forecasts and projections reported in a trajectory format.

In this study, we define the energy score, outline methods to adapt it for specific applications, and perform synthetic experiments to explore its properties. We then apply the energy score to evaluate the performance of models contributing to the 2023–24 Flu Scenario Modeling Hub (SMH). In the Flu SMH, modeling teams provide predictions about future influenza trajectories under certain assumptions about future human behavior, environmental factors, and circulating strains. The SMH has performed 5 cycles of influenza scenario projections; 3 during the 2022-23 influenza season and 1 for each of the 2023-24 and 2024-25 seasons in addition to 19 scenario projections of COVID-19 and 2 of RSV (Scenario Modeling Hub, 2024b). With its recent transition to trajectory-based submissions, the SMH provides the ideal datasets for demonstrating the utility of the energy score in a real-world context (Howerton et al., 2023; Runge et al., 2024).

Scenario projections typically aim to answer policy questions based on specific assumptions rather than predict as accurately as possible future disease trajectories. Therefore, scenario model evaluation can be complex, and must account for both the accuracy of projections against surveillance data and how well the scenario assumptions compare with reality (Howerton et al., 2023; Runge et al., 2024). This differs from short-term forecasts, where the goal is to predict as accurately as possible future disease outcomes. In practice, scenario projection evaluation often focuses on how well the projected trajectories capture the future dynamics of the epidemic (Loo et al., 2024; Reich et al., 2022; Bay et al., 2024). While other established scoring rules remain valuable and should continue to be used where appropriate, our analysis shows that the energy score is a rigorous metric for evaluating both individual models and ensemble projections, and is particularly well-suited to trajectory-based epidemic predictions. By comparing it with other scoring rules, we illustrate its distinct advantages in capturing the full dynamics of stochastic trajectories and advocate for its adoption among the tools routinely used for epidemic model evaluation.

## Methods

When evaluating probabilistic predictions, it is important to use proper scoring rules for model evaluation. Proper scoring rules are evaluation measures such that a forecaster has no incentive to predict anything other than their own true belief (Bracher et al., 2021; Gneiting and Raftery, 2007). If *G* is the underlying generative process of the observations **y**, the score comparing the observed data with *G* will on average give the optimal score. Let *S* (*P*) ≡ *E*_**y**∼*G*_[*S* (*P*, **y**)] be the expected score of a stochastic process *P* with respect to the generative process of the observations *G* (Gneiting et al., 2008). *A scoring rule is strictly proper if the generative process of the observations G* gives the best (minimal) expected score, and any prediction *P* will produce a greater expected score unless *P* = *G*. This can be shown by

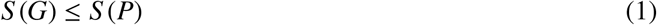

and is equal only if *P* = *G*. This inequality holds for negatively-oriented scores, meaning that smaller scores are considered better predictions. A score is proper but not strictly proper if the inequality holds, but is not uniquely minimized by the generative process *G*. This means that a prediction could give an optimal score even if it is not identical to the generative process of the observed data. The energy score is a strictly proper negatively-oriented score and it is the multivariate generalization of the continuous ranked probability score (CRPS) (Gneiting and Raftery, 2007; Gneiting et al., 2008).

### The Energy Score for Trajectory-based Projections

The concept of the energy score is derived from energy statistics and it measures distances between statistical observations to quantify differences between distributions(Székely and Rizzo, 2017; Székely and Rizzo, 2013). The energy score (ES(*P*, **y**)) of a multivariate distribution *P*, where **X**(*i*) and **X**(*j*) are vectors describing realizations of random variables drawn from *P*, and **y** is the vector of observed values, is defined as:

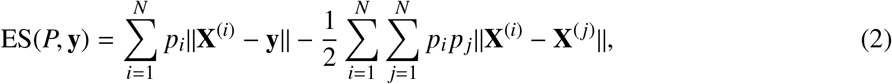

where ∥ · ∥ is the Euclidean norm, *p*_*i*_ is the weight attributed to each individual trajectory *i*, and *N* is the total number of trajectories being analyzed (Gneiting and Raftery, 2007; Staid et al., 2017). The weights *p*_*i*_ can be defined such that trajectories with a higher probability of occurrence are given more weight in the evaluation (Staid et al., 2017). This approach could be applied when evaluating an ensemble model composed of multiple individual models or scenarios grouped together, where trajectories from certain models or scenarios are expected to perform better than others. In the following, we will assume that in projections from a single model, all trajectories are weighted equally, as changing the weights affects the interpretation of the energy score. If we assume that all trajectories are equally weighted with weight 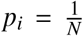, and we expand the Euclidean norm, then Eq. 2 can be written as:

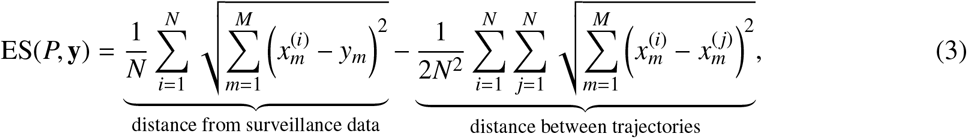

where *M* is the number of elements in each vector (i.e. the number of time points projected), *x*^(*i*)^ is the predicted value by trajectory *i* at time *m*, and *y*_*m*_ is the value of the surveillance data at time *m*. In this definition, the trajectory could be a time-series, where each entry is a prediction for a given date, or more generally a vector of predictions, such as different outcome targets. As we will discuss later, the score is composed of two components, the first term which compares the distance between the predicted trajectories and the observed data, and the second reflects the distances of the trajectories to each other.

The energy score in Eq. 3 is an absolute measure, meaning it is strongly influenced by deviations from the signal at higher magnitudes. In other words, if a trajectory has the same relative error for two observed values of different magnitudes, the energy score will place significantly greater weight on the deviation associated with the larger magnitude. This can be a desirable feature in scenarios where errors on large data points have more serious implications. For instance, a 30% relative error in hospitalization at the onset of an epidemic might correspond to only a few weekly admissions, while the same error at the peak of the season could represent a significant underestimation of hospital bed demand. In such cases, the energy score appropriately penalizes deviations at the epidemic peak more than those at the tail (Bracher et al., 2021; Cramer et al., 2022b).

However, in other situations, this feature may be less desirable—for example, when comparing forecast quality across states with inherently different epidemic curve magnitudes due to varying population sizes. In such cases, a relative energy score can be defined by normalizing the score by the sum of the observed time series data, thereby rescaling the score by the signal’s overall size (Gneiting et al., 2008). A normalized energy score can be written as:

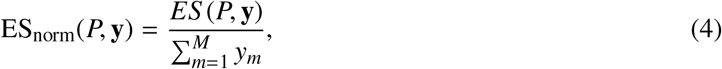

which provides a relative measure that facilitates comparisons of normalized energy scores across different locations. This normalization still emphasizes deviations at the peak within each state but adjusts the overall score based on the sum of the signal, enabling fairer comparisons between states with different population and outbreak sizes (Cramer et al., 2022b). This approach can also be extended to compare scores across different projection targets or time periods. Other normalization methods, typically applied to the WIS, have been proposed for epidemic model evaluation. These include a logarithmic transformation of scales (Bosse et al., 2023), rescaling by the standard deviation across models (Howerton et al., 2023), and pairwise model comparison using a geometric mean (Cramer et al., 2022b). While these methods can be applied to the energy score, we choose to normalize by the surveillance data magnitude as this approach maintains importance of the epidemic peak and avoids relying on other model projections. The issue of normalization is similarly relevant in the multi-dimensional extension of the energy score, as discussed in the next section. Unless otherwise specified, we will use the energy score as defined in Eq. 3.

### Multi-Dimensional Energy Score

One benefit of the energy score is that it can be adapted into a performance measure across multiple dimensions. This would allow us to evaluate a model across multiple target outcomes (i.e. cases, deaths, or hospitalizations), age groups, locations, and so on, with a single score, giving us a comprehensive understanding of a model’s performance with respect to all of its predictions in high-dimensional space. With other scoring rules, this would only be possible via a summary statistic, such as a sum or average of scores for each outcome variable. To calculate this, we look at each time point as a multi-dimensional vector, where *T* represents the number of prediction targets we are assessing. Now, we have a matrix where the columns describe the predictions at each time point, and the rows show the time-series predictions for each outcome. In this definition of the energy score, we are looking at the distance between matrices instead of the distance between vectors, as in the standard energy score. Therefore, we can use the Frobenius norm to measure distance, as it is the multi-dimensional extension of the Euclidean norm, and define the multi-dimensional energy score as:

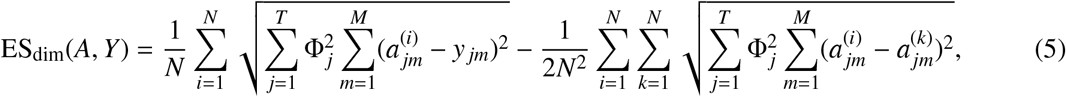

where *A*^(*i*)^ is a *T* x*M* matrix of predictions for trajectory *i*, with *T* outcomes (i.e. prediction dimensions) and *M* time points, Φ _*j*_ is a normalization factor for the magnitude of the signal along each dimension/target, and *N* is the number of trajectories reported for each outcome target. The *T* x*M* matrix *Y* describes the surveillance data, matching the construction of the trajectory matrices *A*^(*i*)^.

The factor 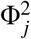 rescales the signal, adjusting the contribution of each forecast target *T* to the multi-dimensional energy score. When 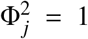, targets with larger magnitudes dominate the score, implying that they are more relevant for assessing model performance. However, this assumption may not always be desirable. For example, if the targets are hospitalizations and deaths, the *T* = 2 energy score will be heavily influenced by the hospitalization target, which typically has a much larger magnitude than the death target. However, it may be preferable for the model to predict both targets with equal accuracy. In such cases, we can use a rescaling factor 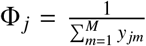, dividing by the sum of the observation vectors for each target outcome to ensure that all outcome dimensions contribute similarly to the multi-dimensional energy score. The same principle applies when forecast targets correspond to different geographical locations. For instance, if each U.S. state is treated as a forecast target, *T* = 50, then the energy score will be dominated by states with larger populations, which typically have higher hospitalization or death counts. Applying a rescaling factor in this context ensures that performance across all states is weighted equally, regardless of population size. The choice of 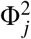 can be adapted on a case-by-case basis, depending on the objectives of the energy score assessment and the desired balance between different forecast targets.

Finally, when constructing the multi-dimensional trajectory matrices, trajectories must be paired appropriately across dimensions. This means that trajectories of different targets (*T*) from the same simulation (*i*) must be combined into the same *T* x*M* matrix *A*^(*i*)^ to preserve correlations between dimensions, as the construction of the time series matrix directly influences the energy score value. There can be cases where the trajectories across dimension do not originate from the same simulation trajectory. When trajectories for each target are generated independently across dimensions, any pairing is acceptable since there are no cross-dimensional correlations. We discuss these cases further in the Results and Supplementary Material. Regardless of the generation method, all matrices must have the same dimensions (i.e. there must be the same number of trajectories for each target). For example, with 100 trajectories in dimension 1 and 200 in dimension 2, only 100 trajectory matrices can be constructed.

## Results

### Comparison of proper vs. strictly proper scoring rules

Several scoring rules have been proposed in the literature to evaluate the performance of probabilistic epidemic projections. Among these, the weighted interval score (WIS) has emerged as a widely adopted standard in forecasting and scenario modeling efforts (Bracher et al., 2021; Howerton et al., 2023). The WIS is a negatively-oriented proper score applied to prediction intervals given by a model’s quantiles. The score consists of three terms that describe the width or uncertainty in the prediction interval, and a penalty if the surveillance data lies outside the prediction interval. It is computed at each time point with prediction *P* and observed value *y* as a weighted sum of the interval score for each prediction interval of interest, and approximates the CRPS (Bracher et al., 2021). In order to evaluate the performance of a full projection time series using the WIS, we take the average of the WIS calculated at each time point, as this ensures that the score remains proper (Bracher et al., 2021). We further discuss the WIS and CRPS in the Supplementary Material.

The energy score and WIS both evaluate a projection based on its calibration, or the distance between the predicted and observed values, and sharpness, which is the amount of uncertainty given by the prediction. We show in Fig. S1A that the energy score and WIS are similar when evaluating a synthetic predictive distribution at a single time point. However, there are differences between the two metrics that must be considered. Most importantly, the energy score is strictly proper for the full projection time series where the WIS is proper, but not strictly proper. This has implications for how these scores evaluate specific probabilistic predictions. We present proofs for the propriety of the energy score and WIS in the Supplementary Material.

To provide a visual intuition of this difference, we generate trajectories from two stochastic epidemic processes that have the same marginal distribution at each time step. First, we generate trajectories from a stochastic SIR model using a chain binomial process. We parameterize the SIR model such that the transmissibility is β = 0.625 and the recovery rate is µ = 0.25 giving us a reproduction number *R*_0_ = 2.5. For the second process, we create a noisy SIR model, where we randomly shuffle the values for the number of infectious individuals at each time point in the SIR model trajectories. Then we randomly group these values across time to produce noisy epidemic trajectories. This gives us two epidemic-like processes with the same marginal distribution at each time step, one with time-correlated, and one with time-uncorrelated stochastic trajectories. An example of the two trajectory processes are shown in Fig. 1A and 1B, where we also show the quantile format for these trajectories in Fig. 1C, which is identical for each trajectory process. We then score the two processes using as observation vector a single realization of the stochastic SIR model described above. Using this framework, we calculate the energy score and WIS for 200 iterations, generating 100 trajectories at 60 time points for each model at every iteration.

**Figure 1:**
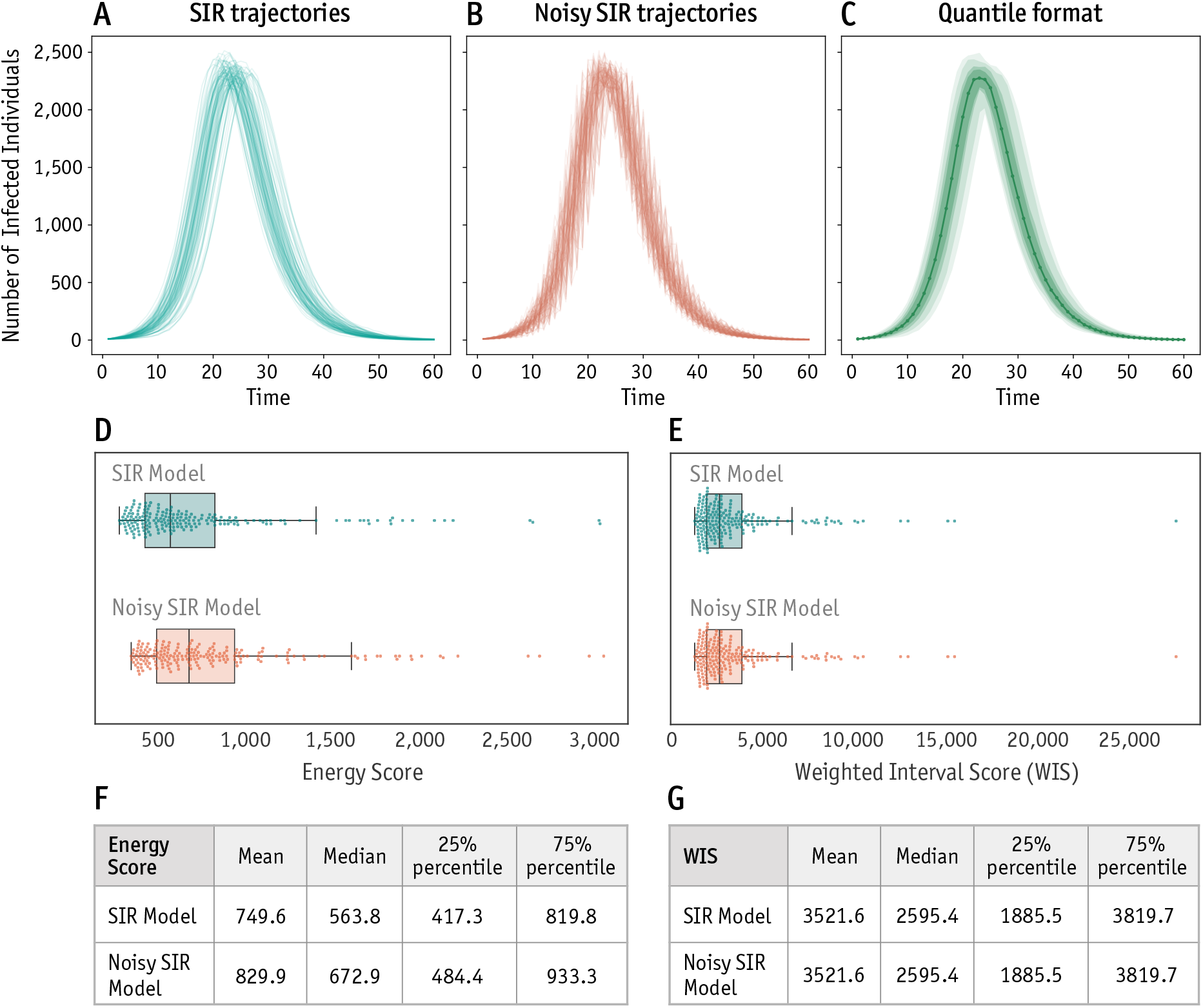
Synthetic experiments comparing energy score and WIS propriety and evaluation. Comparison of the WIS and energy score for (A) SIR model and (B) noisy SIR epidemic trajectory process with the same marginal distribution at each time point, generating the same quantile format (C) with median (solid line) and 50%, 70%, 90%, and 98% prediction intervals shaded. Boxplot of the energy score (D) and WIS (E) obtained from each trajectory process given 100 trajectories each, showing the distribution for each score and epidemic trajectory process across 200 iterations. For visualization purposes, the maximum values for each score and model type is not shown. Boxplots are created such that the box shows the 25%, 50% and 75% quantiles, and the whiskers represent 1.5× interquartile range (IQR). Descriptive statistics of the (F) energy score and (G) WIS for the SIR and noisy SIR model.

In Fig. 1D and E, we show the distribution of the scores for each epidemic trajectory process, and descriptive statistics of these distributions in Fig. 1F and G. From the boxplots and tables of descriptive statistics, we find that the WIS for both model processes is the same, but the energy scores differ. The energy score is able to distinguish between two processes with the same marginal distribution but differing individual behavior, where the WIS scores them identically. The SIR model process, on average, produced better values of the energy score than the noisy model, which agrees with the knowledge that the observed values were generated from the SIR model. This difference is due to the energy score being strictly proper, while the WIS is proper but not strictly proper. If a prediction *P* has the same marginal distribution as the true underlying process *G*, it would give the ideal WIS score to both even if *P* * *G*; on the other hand since the energy score is strictly proper, only a prediction *P* = *G* can give the ideal energy score (Gneiting and Raftery, 2007). Moreover, the fact that on average the energy score gives lower scores to the SIR model demonstrates the impact that the additional information contained within individual trajectories provides. Since the noisy SIR model trajectories do not follow the same temporally correlated behavior as the SIR model, and therefore the observation vector, they are penalized.

### Assessing Forecast Models with Multiple Peak Timing: Advantages of the Energy Score

A fundamental distinction between the energy score and the WIS lies in how they capture the epidemic dynamics. While the energy score evaluates the complete set of individual trajectories directly, the WIS relies on summary statistics, typically quantiles, computed at each fixed time point. This difference has important implications for how well each scoring rule captures key dynamic features of epidemic forecasts, such as the timing and magnitude of epidemic peaks.

Fixed-time quantile summaries, as used in the WIS, tend to obscure multi-modal or non-synchronous behaviors across trajectories. For example, consider a model in which each trajectory includes a single epidemic peak, but the peak time of the trajectories vary. When trajectories are aggregated into quantiles at each time point, the resulting summary can flatten or even mask these peaks entirely. The quantile representation may suggest a smoother, less peaked epidemic curve that does not correspond to any individual realization of the model (Juul et al., 2021).

This effect is especially problematic in the context of scenario modeling. With scenarios, models should not only predict the most likely outcome, but also explore plausible alternative trajectories, including worst-case outcomes. When such forecasts are scored using the WIS, models that correctly capture complex behaviors, such as epidemics having a wide distribution of peak time, may be unfairly penalized if their median or central quantile predictions deviate from the observed data, even if a substantial subset of trajectories match the observed epidemic.

To illustrate this limitation, we designed a synthetic forecasting experiment involving the superposition of two stochastic SIR models, each producing a single epidemic peak at a different time. We generate a standard stochastic SIR model (as described in the previous section) with low variability in the peak timing. Additionally, we generate trajectories from a double-peak SIR model with trajectories having a different peak distribution: each realization contains a single peak, but across the ensemble, peaks occur at two distinct time points. Fig. 2A shows the trajectories and corresponding quantile representations of the single peak and double-peak SIR models. For evaluation, we selected a single realization of the single peak SIR model as the ground truth, and we compare forecast scores using both the energy score and the WIS. For the WIS calculation, quantiles were estimated from the trajectories generated by the SIR models. We repeat this process generating 100 trajectories and scoring each model for 500 iterations, and test different widths between the two peaks in the trajectories, namely peaks that are separated by 5, 10, or 20 time steps.

**Figure 2:**
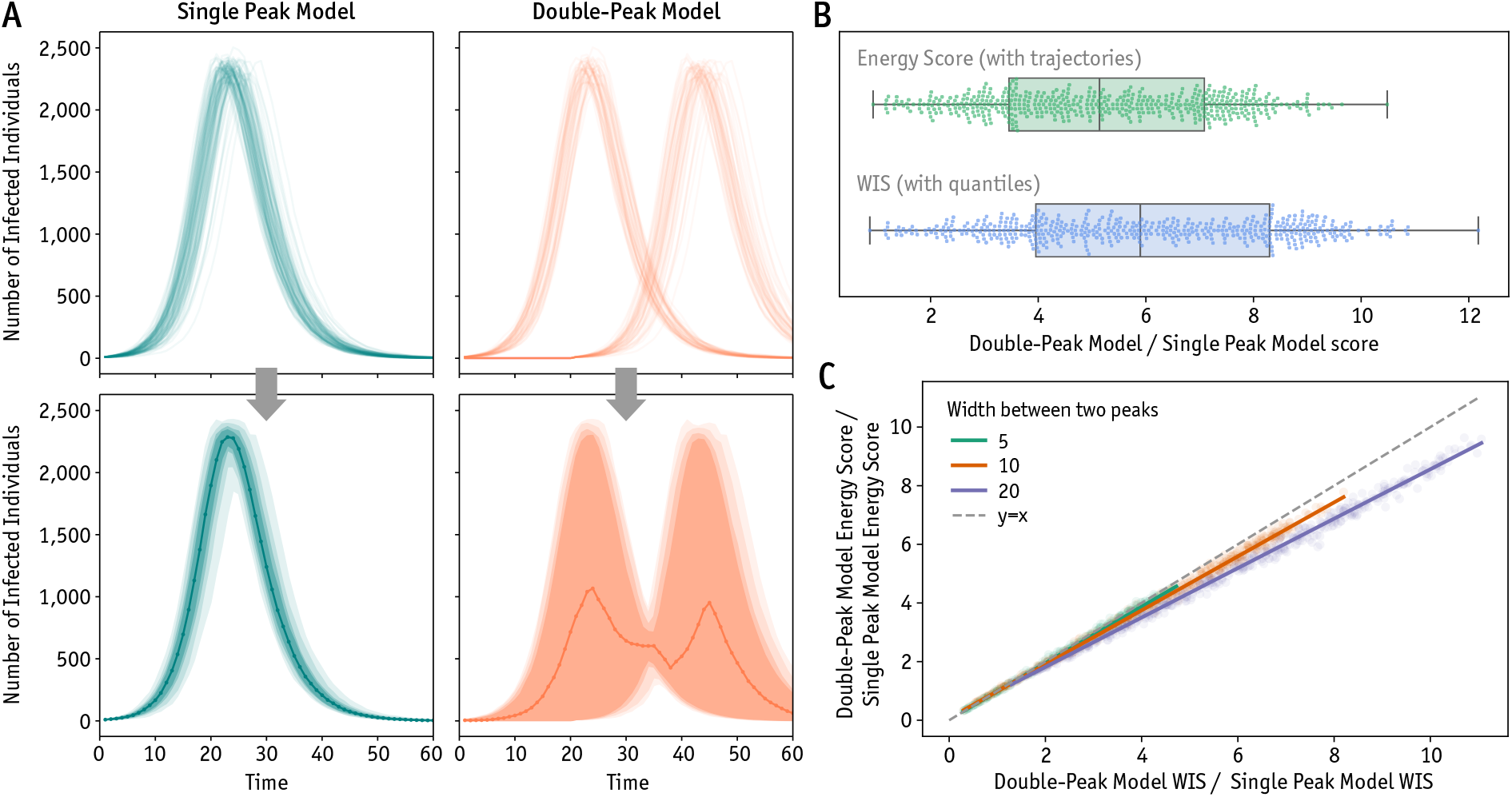
(A) Single peak (left) and double-peak (right) stochastic SIR model trajectories (top) and corresponding quantile format for the 50% and 98% prediction interval (bottom). Shown for a width between the two peaks of the trajectories of 20 time steps. (B) Distribution of the ratio between double-peak model and single peak model score for the energy score and WIS for 500 iterations of generating 100 trajectories for each model, with a distance of 20 time steps between the 2 peaks in the double-peak model’s trajectories. Boxplots are such that the box shows the 25%, 50% and 75% quantiles, and the whiskers represent 1.5× interquartile range (IQR). (C) Relationship between the double-peak model / single peak model score ratio for the energy score and WIS for different distances between the two peaks in the model. Dashed gray line shows y=x and the solid lines show the linear regression fits of the data for the different widths between peaks. Dots show the scores for each iteration of trajectory generation and scoring.

We compare the behavior of the two scores by finding the ratio between each score given by the double-peak model divided by the corresponding score for the single peak model. As shown in Fig. 2B, the forecast model with the double-peak structure is significantly more penalized by the WIS than by the energy score. The ratio between the double-peak model and single peak model are typically lower for the energy score than the WIS. This discrepancy increases with the growing separation between the two peaks, as confirmed by our correlation analysis of the relative scores in Fig. 2C. As the two peaks grow farther apart, the ratios for the energy score and WIS become increasingly different. The energy score, which directly evaluates the distribution of complete trajectories, is able to identify subsets of realizations that accurately match the observed dynamics.

These findings indicate a potential limitation of quantile-based evaluations in capturing complex epidemic behaviors. When model outputs display substantial peak-timing uncertainty or multi-modal epidemic dynamics, the energy score offers a more appropriate measure of predictive performance, as it directly evaluates the full set of stochastic trajectories. This property makes it particularly well-suited for assessing forecasts and projections reported in trajectory format. Figure S2 illustrates how similarly complex dynamics can lead to marked differences in scoring during real-time epidemic projection exercises.

### Case Study from the Flu Scenario Modeling Hub

We illustrate the application of the energy score to epidemic scenario projections for the 2023-24 projection round of the Flu Scenario Modeling Hub. In previous projection rounds, modeling teams were only required to report quantiles of their predictions aggregated for each week, but beginning in the 2023-24 influenza round, teams submitted at least 100 individual trajectories from their model output for each scenario, and location (Loo et al., 2024). The objective of this projection round was to explore the implications of different vaccine coverage levels (high, normal, or low) and the dominant circulating strain (either A/H3N2 or A/H1N1) on the trajectory of weekly hospitalizations during the 2023-2024 flu season for US states and nationally (Scenario Modeling Hub, 2024a). Modeling teams reported projections for these 6 scenarios, from September 3, 2023 to June 1, 2024. Further information about the projection round and model output can be found in the Flu SMH’s GitHub repository. Table S1 describes the 2023-24 SMH scenarios in further detail. In Fig. 3, we show the reported model output for the ‘MOBS_NEU-GLEAM_FLU’ model in the US nationally for all 6 scenarios of the 2023-24 Flu SMH round. We see how the scenarios result in distinct epidemic outcomes given assumptions on the circulating influenza strain and vaccine uptake. We analyze the incident hospitalization projections from September 9, 2023 to April 27, 2024.

**Figure 3:**
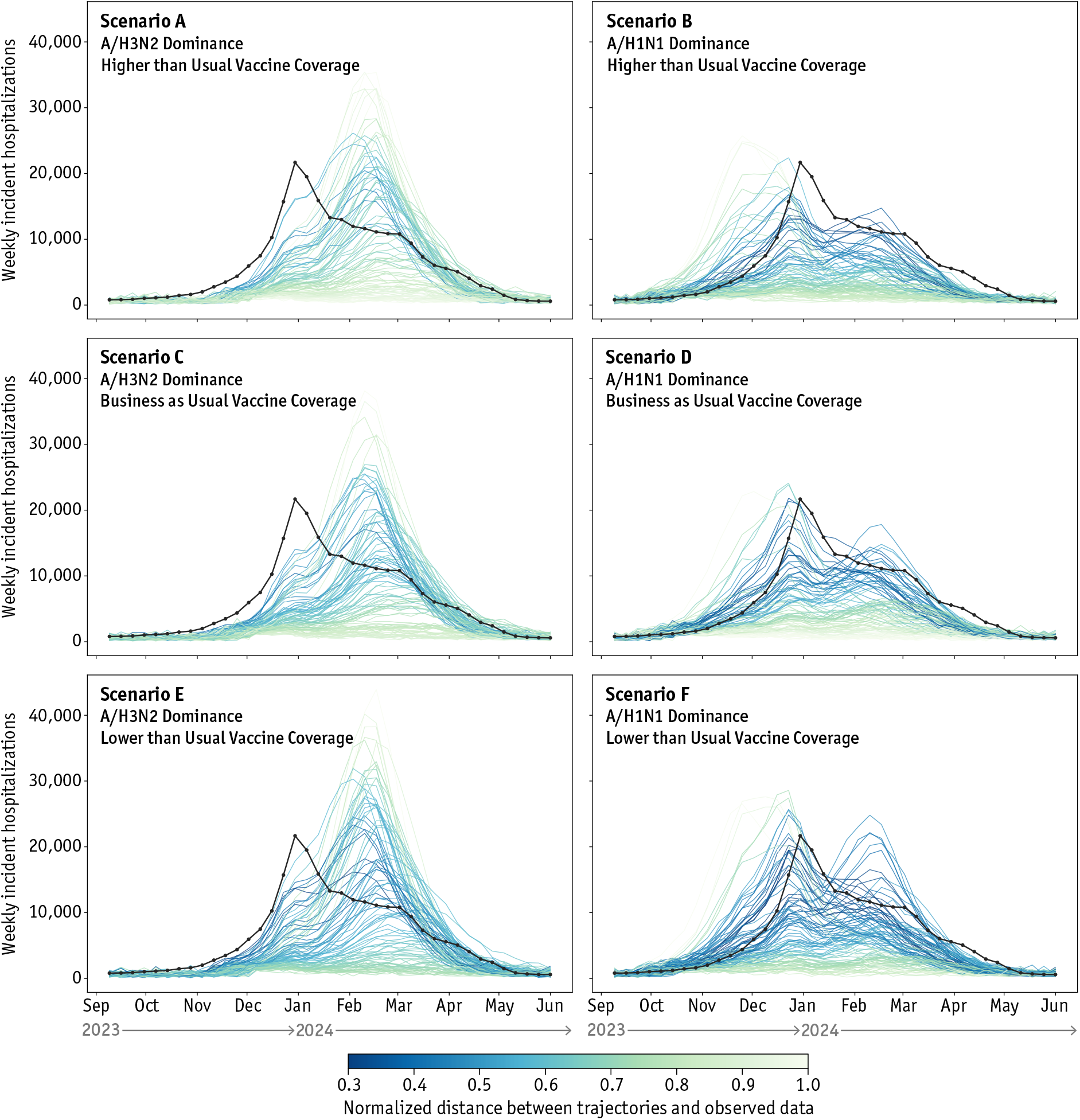
Epidemic predictions in the trajectory format. One hundred trajectories for incident hospitalizations for the ‘MOBS NEUGLEAM FLU’ model for all 6 scenarios in the Flu Scenario Modeling Hub 2023-24 round 1 with observed surveillance data (black) nationally in the United States. Trajectories are colored by their normalized distance from the observed data. Lighter colors represent a larger distance.

Given the widespread use of the weighted interval score (WIS) for evaluating probabilistic epidemic predictions, we compare the performance assessment provided by the WIS to that of the energy score to understand how these metrics evaluate model projections. We further describe how the WIS is computed from the Flu SMH projections in the Supplementary Material.

### Single Model Evaluation

We focus the first part of our analysis on the ‘MOBS_NEU-GLEAM_FLU’ model submitted to the Flu Scenario Modeling Hub, which is a multi-scale, age-structured, stochastic metapopulation epidemic model that uses global flight and commuting data to simulate the spread of an infectious disease (Balcan et al., 2010; Chinazzi et al., 2024). In this section, we examine the energy score for this single model across time periods, locations, and scenarios.

As noted earlier, the energy score is influenced by the magnitude of values defined in each trajectory. This dependency complicates the comparison of scores across locations, time periods, or target outcomes with differing magnitudes. In Fig. 4A, we compare the energy score for the ‘MOBS_NEU-GLEAM_FLU’ model influenza hospitalization projections for each scenario and location to the sum of the corresponding surveillance time series. We find that the non-normalized energy score is strongly correlated to the size of the signal. However, Fig. 4B shows that the normalized energy score depends much less on the magnitude of the surveillance data, with an *R*^2^ of 0.018. This means that scores can be compared with the normalized energy score, and demonstrates its nature as a relative score.

**Figure 4:**
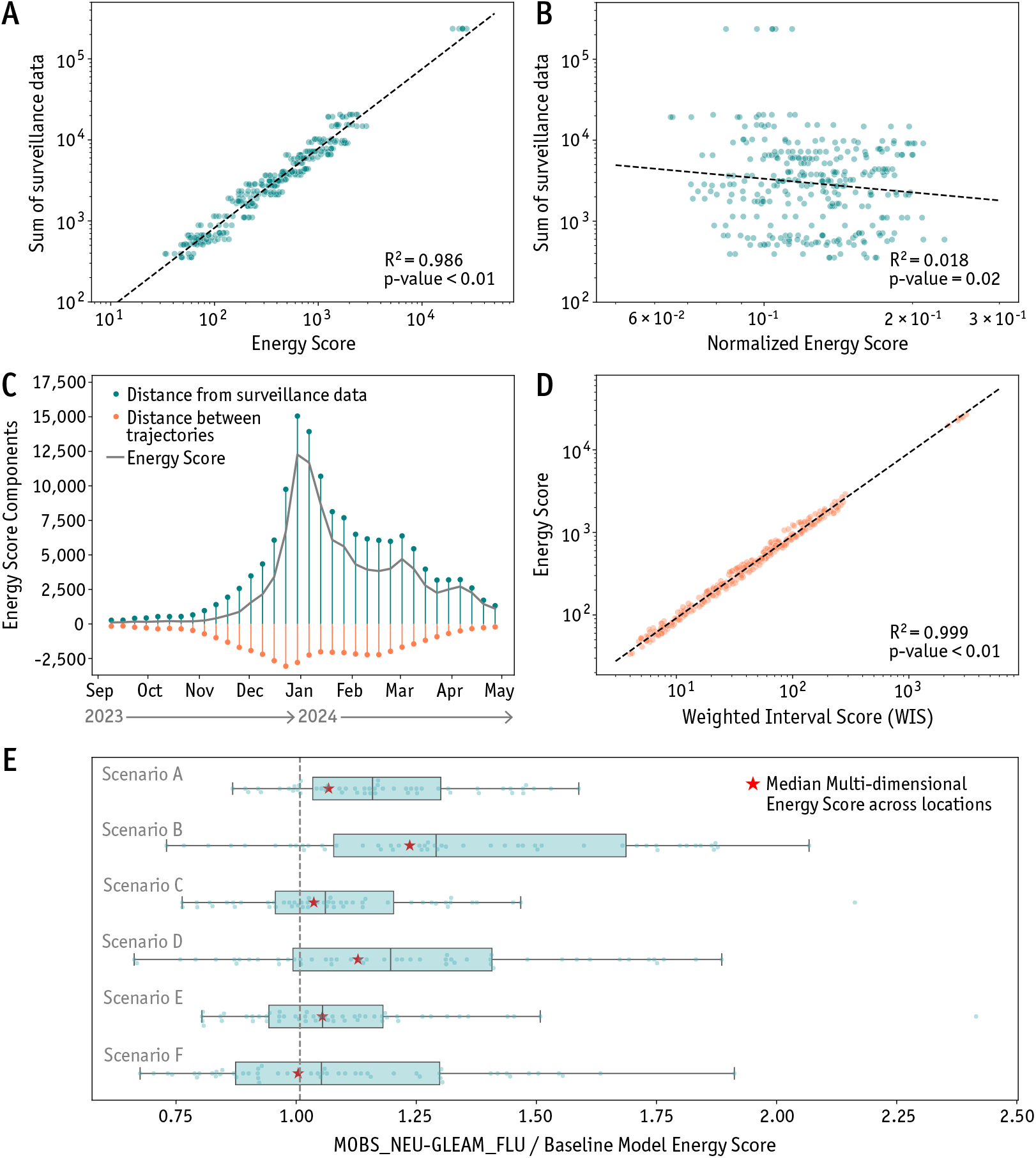
Evaluation of a single model with the energy score. Comparison of the dependence of the (A) energy score and (B) normalized energy score on the sum of the surveillance data of each scenario and location for the model projections. (C) Decomposition of the energy score terms at each week for the Scenario D projections, where the blue represents the term describing the distance of the trajectories from observed data, and the orange describes the term representing the distance between all pairs of trajectories. The gray line shows the full energy score calculated at each week. (D) Relationship between the WIS and energy score. Each point describes the WIS and energy score for a location and scenario. The WIS was found by estimating the quantiles estimated from the trajectories. (E) Boxplot of energy score ratio across 52 locations for the ‘MOBS_NEU-GLEAM_FLU’ model compared to a 4-week-ahead naive baseline model for each scenario. Vertical dashed line shows where the ‘MOBS_NEU-GLEAM_FLU’ and baseline model have the same scores, where ratios below one describe when the model performs better than the baseline. The overlaid red stars show the median of the multi-dimensional energy score ratio across locations for 50 iterations of randomizing the trajectory pairings. Boxplots are created such that the box shows the 25%, 50% and 75% quantiles, and the whiskers represent 1.5× IQR. Scatter plots (A,B,D) show a dashed line of the linear regression fit, with the *R*^2^ and two-sided p-value describing the fit. All results are shown for the ‘MOBS_NEU-GLEAM_FLU’ model 2023-24 Flu SMH incident hospitalization projections.

While the energy score is defined for a multivariate time series, we analyze its behavior at individual time points to study how the energy score changes throughout an outbreak. In Fig. 4C, we show the energy score at each week of the flu season (Eq. 3 where M=1) for the ‘MOBS_NEU-GLEAM_FLU’ model, scoring incident hospitalization projections for Scenario D (A/H1N1 Dominance, Business as usual vaccination coverage) nationally in the U.S., and we decompose these scores to look at the contribution of individual terms. The energy score is made up of a positive term describing the distance between the trajectories and surveillance data, and the pairwise distance between all trajectories subtracted from this value. This negative term is why the energy score value is less than the component describing the distance from surveillance data. In this example, the energy score is largely composed of the distance between each trajectory and the observed data. The score increases at time points near the peak as expected because the energy score is an absolute metric depending on the magnitude of the surveillance signal, which means it will typically give higher weights near the peak of an epidemic curve. We show in Fig. S1B that the WIS follows similar patterns when evaluated at each week of the influenza season.

Since the WIS is a commonly used score for evaluating epidemic projections, we compare the performance of the ‘MOBS_NEU-GLEAM_FLU’ model using the energy score and WIS. If we calculate the energy score and WIS for the predictions made for each location and scenario for incident hospitalizations in the 2023-24 Flu Scenario Modeling Hub round, we find a strong correlation between the two scores, shown in Fig. 4D. This highlights that a prediction that performs well under the WIS is likely to also be scored well by the energy score. In Fig. S1C, we show a strong correlation between the normalized energy score and normalized WIS as well.

Using the energy score, we are also able to compare performance across scenarios and locations. In Fig. 4E, we evaluate the performance for all scenarios, looking at the distribution of scores for predictions at each location. To evaluate model performance, we create a naive baseline forecast for 4-week-ahead projections, to use as a reference point against which we compare the performance of long-term scenario projections (Ray et al., 2023). While alternative baseline methods can be devised for scenarios projections, this approach has been used previously to assess the performance of several rounds of the SMH COVID-19 projections (Howerton et al., 2023). The baseline forecast generates projections where the median is equal to the most recently observed data point, with expanding uncertainty for longer horizons informed by historical changes in incidence counts for consecutive weeks. We report additional details on the generation of the naive baseline model and its trajectories in Fig. S3 in the Supplementary Material. From the baseline model, we create an energy score ratio where we divide the energy score for each model, location, and scenario by the energy score of the baseline model for each location. A ratio less than 1 describes when the scenario model performs better than the naive baseline model. It is important to note that scenario projections are not necessarily expected to outperform the baseline model. This is because the baseline model is continuously updated with new data each week to produce 4-week-ahead predictions, whereas scenario projections are made several months in advance without incorporating recent surveillance data. In our analysis, we find that most scenarios for the ‘MOBS_NEU-GLEAM_FLU’ model perform similar to or slightly worse than the naive baseline model. In Table S2, we show descriptive statistics for the energy score ratio ‘MOBS NEUGLEAM FLU’ model for each scenario.

We also utilize the multi-dimensional energy score to compute a single comprehensive score for each scenario, providing an overall assessment of the ‘MOBS_NEU-GLEAM_FLU’ model’s performance across all U.S. states. The multi-dimensional energy score requires trajectories to be paired consistently across all dimensions, including across states. However, the SMH does not explicitly provide the pairing of trajectories across different locations. For the ‘MOBS_NEU-GLEAM_FLU’ model, trajectories in different states are originated from independent calibrations, allowing us to randomize the trajectory identifiers and conduct a sensitivity analysis to evaluate how different pairings influence the multi-dimensional energy score, repeating this process over 50 iterations. Additionally, we calculate an energy score ratio, as previously described, to compare these values against the multi-dimensional energy score of the 4-week-ahead naive baseline model. In Fig. 4E, the red stars show the median multi-dimensional energy score ratio across these iterations for each scenario, where the dimensions are U.S. states. In Table S5, we show that the uncertainty around the multi-dimensional energy score for randomizing the trajectory pairings is very small. Additionally, we find that the distribution of the energy score ratio across locations in the boxplots and the multi-dimensional energy score rank the scenarios similarly, with both medians following the same pattern. This shows that the multi-dimensional energy score is evaluating the model similarly to the standard energy score, but in an aggregated manner. In addition, we examine the impact of removing the normalization factor from the multi-dimensional energy score expression in order to keep emphasis on locations with larger signals in Fig. S4.

### Multi-Model Analysis

In this section, we focus our analysis on the performance of all models submitted to this Flu SMH projection round that provided incident hospitalization projections for multiple locations. This selection resulted in six individual models being included in the analysis, with four models excluded from the study because they only reported data for one location. Ensemble models generated by the SMH were not considered, as these models report results only in a quantile format. To illustrate the utility of the energy score in comparing performance across multiple models, we analyze model rankings and performance across different locations. In Fig. 5 we show how the energy score varies across models, scenarios, and locations. We calculate the energy score ratio in comparison with the 4-week-ahead naive baseline model (Ray et al., 2023) described previously for each scenario, location, and model, where a ratio below one represents a case where the scenario model performed better than the naive baseline model. We find that there are models that consistently perform better than others, and that some models perform very similar to or slightly better than the baseline model. Table S3 presents descriptive statistics of this data for all scenarios and models by summarizing the energy score ratio for all models and scenarios examined. This demonstrates the ability of the energy score to compare models and differentiate between better-performing models. This tells us about relative model performance within each scenario.

**Figure 5:**
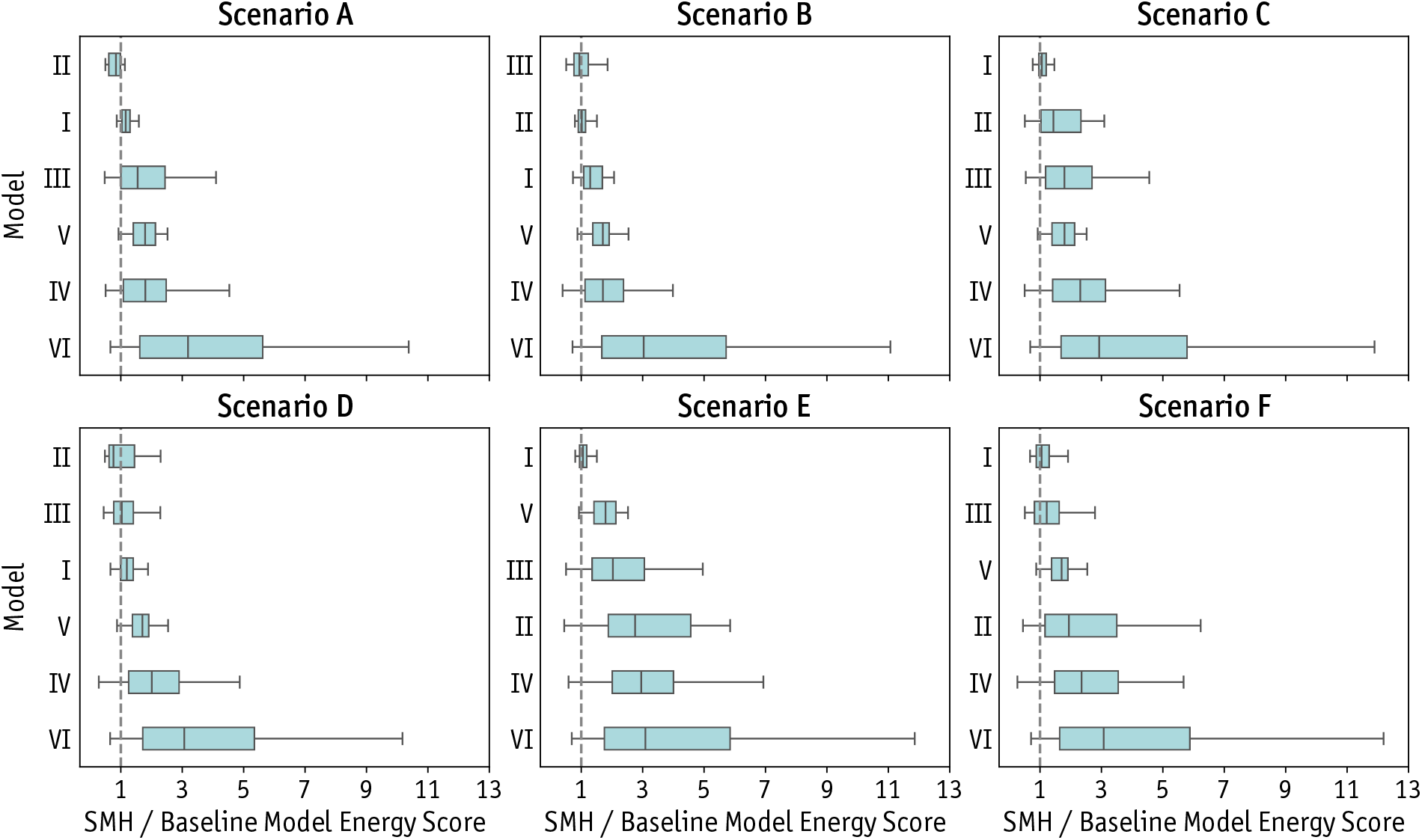
Energy score ranking across models. Boxplots showing the distribution of the ratio of the energy score for each model, location, and scenario in Flu Scenario Modeling Hub divided by the energy score for the 4 week ahead naive baseline flu model at each location. Each boxplot shows the energy score ratio distribution over the number of locations the corresponding model reported (shown in Table S3). Vertical dashed line shows where the ensemble and baseline model has equal energy scores such that ratios below one describe when the SMH model performs better than the baseline. Models are ordered by median energy score ratio within each scenario. Boxplots are such that the box shows the 25%, 50% and 75% quantiles, and the whiskers represent 1.5 × IQR. Note that Model I represents the ‘MOBS_NEU-GLEAM_FLU’ model, which can be compared to results in Fig. 4.

We also investigate how model rankings based on the energy score compare to those derived from the WIS. For each scenario and location, we rank the models from lowest to highest score using both the energy score and WIS. To assess the agreement between these rankings, we calculate Kendall’s τ rank correlation across all models. The Kendall’s τ correlation coefficient compares rankings by counting the number of pairs of objects that are in an incorrect order, divided by the total possible pairs (Kendall, 1938). A rank correlation of 1 means that the rankings of the energy score and WIS are identical. In Fig. S1D, we show a histogram of the Kendall’s τ rank correlation between the energy score and WIS for all models and scenarios. The mean rank correlation coefficient between the energy score and WIS is 0.87. More precisely, we find that for 49.7% of projections, the energy score and WIS have a Kendall’s τ correlation coefficient of 1. This shows that the energy score and WIS rank models quite similarly, but in a majority of cases, the rankings are not identical.

### Trajectory-Based Ensemble

In multi-model analysis, it is common practice to create an ensemble model by combining predictions from the multiple individual models. (Ray et al., 2023; Reich et al., 2022). In the ensemble, the information and uncertainty from multiple models are aggregated together to provide a “consensus” projection of future possibilities aggregating the different assumptions and methodologies of individual models (Cramer et al., 2022b). The performance of ensemble models has been shown to generate improved future predictions that characterize uncertainty better than individual models (Bates and Granger, 1969; Cramer et al., 2022b; McGowan et al., 2019; Reich et al., 2022). The SMH includes three ensemble models as part of its analysis. However, these ensembles are built by first converting each model’s trajectories into a quantile format before applying the ensemble methods. Here, we propose an alternative approach for generating an ensemble model that directly utilizes the trajectories reported by each modeling team. In this method, we simply bundle all trajectories from each model into the definition of a single ensemble, assigning equal weight to each trajectory. In our case, each model contributes the same number of trajectories. If this is not the case, weighting or sampling methods could be considered so that models reporting more trajectories do not have an disproportionate influence on the ensemble behavior. This ensemble approach avoids the need to summarize stochastic model outputs into quantile format, leveraging the raw data provided by the models instead. In Fig. S5, we compare this ensemble of trajectories method with the three ensemble models reported by the SMH using the WIS. We cannot calculate the energy score of the SMH-reported ensemble models because they are only reported in quantile format, so trajectories are not available.

In Fig. 6, we explore the energy score across locations using the trajectory-based ensemble. We include all models reported to the 2023-24 Flu SMH that submit projections of incident hospitalizations for any number of the 50 U.S. states, Washington D.C., or nationally. Fig. 6A shows the ratio of the energy score of the trajectory-based ensemble model divided by the energy score of the 4-week-ahead naive baseline model at each location (U.S. states and the District of Columbia) for Scenario D. Locations in blue show where the ensemble model performed better than the baseline model (ratio < 1), where those in orange show where the naive baseline model had better performance (ratio > 1). We observe heterogeneous performance across the United States, with the ensemble model outperforming the naive baseline in 24 locations, but falling short in 27. These results indicate areas where model performance could be improved for future predictions. Notably, in this example, the ensemble tends to perform better in the Southern U.S. states.

**Figure 6:**
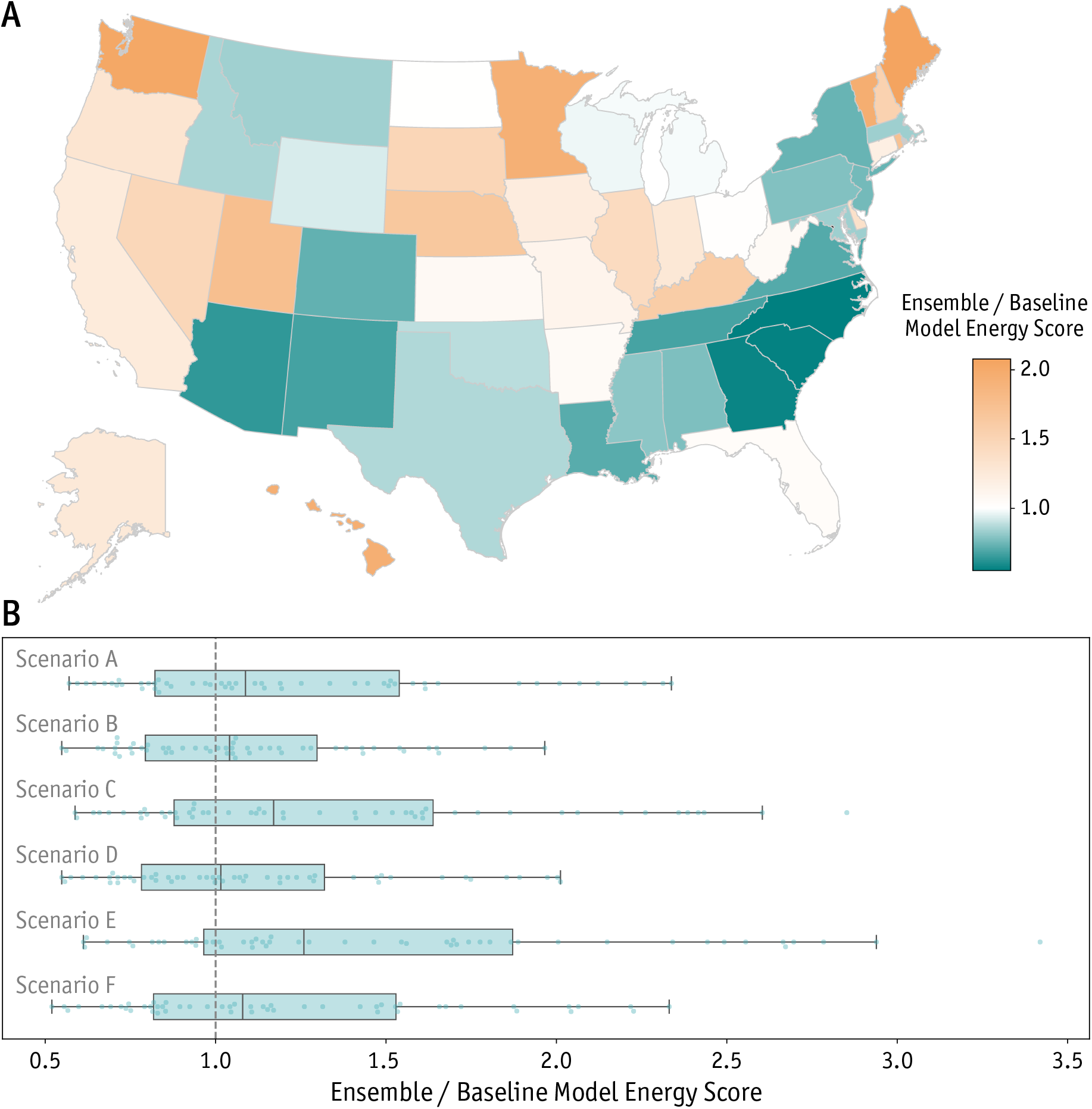
Energy score for the trajectory-based ensemble model. (A) Map of the US showing the energy score ratio (trajectory-based ensemble / baseline model energy score) for each US state of the 2023-24 Flu Scenario Modeling Hub round for Scenario D. Ratios below one (blue) show where the energy score for the trajectory-based ensemble model for a state was better than the naive baseline, where the ratios above one (orange) show where the baseline model had better performance. (B) Boxplots of the energy score ratio across 52 locations for all scenarios compared to a 4-week-ahead naive baseline model. Vertical dashed line shows where the ensemble and baseline model has equal performance, where ratios below one describe when the ensemble model performs better than the baseline. Boxplots are such that the box shows the 25%, 50% and 75% quantiles, and the whiskers represent 1.5 × IQR.

We extend this to show the distribution in the energy score ratio compared to the 4-week-ahead naive baseline model across locations for each scenario in Fig. 6B. This allows us to assess the overall performance of the trajectory-based ensemble model for each scenario, and compare the scenarios to each other. This analysis illustrates that scenarios B, D, and F are the best-performing scenarios for the ensemble model, which corresponds to the scenarios describing H1N1 as the dominant circulating strain. This is in agreement with the observed strain dynamics in the United States for the 2023-24 influenza season (Centers for Disease Control and Prevention, 2024). We show summary statistics of this data in Table S4.

It is worth noting that we do not calculate the multi-dimensional energy score for the ensemble model due to the assumptions required about trajectory independence and pairing—information that is unavailable for the other modeling teams within the SMH. The multi-dimensional energy score relies on constructing matrices that group trajectories across dimensions, so unknown dependencies could distort the score by introducing unexpected dependencies. Ignoring these assumptions could bias the multi-dimensional energy score and compromise the results of these calculations.

This ensemble of trajectories method could be easily translated to a different weighting process where models or even specific trajectories are weighted differently as data is gathered (Sherratt et al., 2024). This allows for a flexible framework for ensemble creation and performance analysis using only single trajectories generated by individual models without the need to summarize and aggregate model output using quantiles.

## Discussion

The energy score is not a new metric and has been widely applied to the evaluation of probabilistic forecasts across diverse fields(Gneiting et al., 2008). Yet, its potential in epidemic modeling has remained largely underexplored. The increasing adoption of trajectory-based submissions in epidemic forecasting underscores the need for evaluation methods that can capture their full complexity. In this context, our findings show the value of the energy score for improving how forecasts are assessed, how uncertainty is communicated, and how model outputs are compared across settings.

Using multiple scoring rules to assess predictions is shown to be useful (Pic et al., 2025), and the energy score adds a strictly proper performance measure to the toolbox of probabilistic prediction analysis methods. The energy score also provides a principled framework for evaluating epidemic model outputs in trajectory format, avoiding the need of the additional aggregation step to summarize stochastic realizations into quantiles. By operating directly on individual trajectories, it captures the full temporal structure and variability of the projections, thereby preserving information that would be partially lost in fixed-time descriptive statistics. In addition, the energy score can distinguish between predictions with identical marginal distributions but different underlying generative processes, thus more accurately assessing complex features such as multi-peak epidemic dynamics. To extend its applicability, we introduce a multi-dimensional formulation that enables a comprehensive evaluation of model performance across multiple targets or locations simultaneously.

One limitation of the energy score is the increasing computational cost of the energy score as the number of trajectories increases. The second term in the energy score expression requires the calculation of the pairwise distance between all trajectories. This can become computationally challenging when the number of trajectories is large. We show in Fig. S6 that randomly sampling even a small percent of trajectories gives good agreement with the energy score found through using the full set of stochastic trajectories.

The discrimination ability of a scoring rule illustrates the differences in scores generated by forecasts of differing quality. For example, a scoring rule has low (high) discrimination ability if forecasts with a very different quality result in the same (different) scoring value (Pinson and Tastu, 2013). A proper scoring rule can still have poor discrimination ability. It has been shown that the energy score lacks discrimination ability between forecasts with different correlation structures, but discriminates well between predictions with different means or variances (Alexander et al., 2022; Pinson and Tastu, 2013; Held et al., 2017). Many of these works are aimed at the capability of the energy score to correctly identify the true underlying distribution driving the dynamics. While this is important to consider when employing the energy score for the performance analysis of epidemic forecasts, we believe that it should not limit our use of this tool. The main goal in evaluating epidemic projections and forecasts is not the specification of the true data-generating distribution, but the identification of projections that are closest to reality.

The analysis and results reported here have considered equally weighted trajectories, but the energy score formulation also allows trajectory weights to be individually defined. This flexibility opens the door to scoring strategies that adjust weights based on past performance or prioritize specific outcomes of interest. Although such modifications may shift the interpretation of the score, they could prove valuable for building ensembles or calibrating models as new information about epidemic progression becomes available.

Beyond the case study of the Flu Scenario Modeling Hub, the energy score is broadly applicable to any epidemic forecast or prediction that produces stochastic realizations. As the reporting of trajectories becomes increasingly common, the energy score should be viewed as a widely applicable tool for model evaluation, enabling the full use of information contained within individual trajectories and supporting transparent and informative assessments of epidemic predictions.

## Data Availability

All data produced in the present work are contained in the manuscript.

https://doi.org/10.5281/zenodo.16898960

## Author contributions

Conceptualization: CB, KM, GS, JTD, AV; Investigation: CB, KM, GS, MC, JTD, AV; Methodology: CB, KM, GS, JTD, AV; Software: CB; Supervision: JTD, AV; Writing–original draft: CB, AV; Analyze the results, writing–review & editing the paper: CB, KM, GS, MC, JTD, AV.

## Data and Code Availability

For surveillance data, we use the National Healthcare Safety Network (NHSN) Weekly Hospital Respiratory Data (Centers for Disease Control and Prevention, 2025) for confirmed influenza hospital admissions. We use a formatted version of this data provided by the CDC FluSight Forecast Hub (FluSight Forecast Hub, 2025). Scenario model projections are made available by the Flu Scenario Modeling Hub (Scenario Modeling Hub, 2024a) and in their GitHub repository. Our code for this project is publicly available on Zenodo (Bay, 2025).

## Acknowledgments

We acknowledge support from HHS/CDC 5U01IP0001137, and the cooperative agreement CDC-RFAFT-23-0069 from the CDC’s Center for Forecasting and Outbreak Analytics. The findings and conclusions in this study are those of the authors and do not necessarily represent the official position of the funding agencies, the CDC, or the U.S. Department of Health and Human Services of the United States.

## References

Alexander, C., Coulon, M., Han, Y., Meng, X., 2022. Evaluating the discrimination ability of proper multi-variate scoring rules. Annals of Operations Research doi:10.1007/s10479-022-04611-9.

Balcan, D., Gonçalves, B., Hu, H., Ramasco, J.J., Colizza, V., Vespignani, A., 2010. Modeling the spatial spread of infectious diseases: The global epidemic and mobility computational model. Journal of Computational Science 1, 132–145. URL: https://www.sciencedirect.com/science/article/pii/S1877750310000438, doi:10.1016/j.jocs.2010.07.002.

Bates, J.M., Granger, C.W.J., 1969. The combination of forecasts. J. Oper. Res. Soc. 20, 451–468. URL: https://doi.org/10.1057/jors.1969.103, doi:10.1057/jors.1969.103.

Bay, C., 2025. clarabay/energy-score. URL: https://doi.org/10.5281/zenodo.16898960, doi:10.5281/zenodo.16898960.

Bay, C., St-Onge, G., Davis, J.T., Chinazzi, M., Howerton, E., Lessler, J., Runge, M.C., Shea, K., Truelove, S., Viboud, C., Vespignani, A., 2024. Ensemble2: Scenarios ensembling for communication and performance analysis. Epidemics 46, 100748. URL: https://www.sciencedirect.com/science/article/pii/S1755436524000094, doi:10.1016/j.epidem.2024.100748.

Bonacina, F., Boëlle, P.Y., Colizza, V., Lopez, O., Thomas, M., Poletto, C., 2024. Characterization and forecast of global influenza (sub)type dynamics. medRxiv URL: https://www.medrxiv.org/content/early/2024/08/02/2024.08.01.24311336, doi:10.1101/2024.08.01.24311336.

Bosse, N.I., Abbott, S., Cori, A., van Leeuwen, E., Bracher, J., Funk, S., 2023. Scoring epidemiological forecasts on transformed scales. PLOS Computational Biology 19, 1–23. URL: https://doi.org/10.1371/journal.pcbi.1011393, doi:10.1371/journal.pcbi.1011393.

Bracher, J., Ray, E.L., Gneiting, T., Reich, N.G., 2021. Evaluating epidemic forecasts in an interval format. PLOS Computational Biology 17, 1–15. URL: https://doi.org/10.1371/journal.pcbi.1008618, doi:10.1371/journal.pcbi.1008618.

Centers for Disease Control and Prevention, 2024. Influenza activity in the united states during the 2023–2024 season and composition of the 2024–2025 influenza vaccine. URL: https://www.cdc.gov/flu/whats-new/flu-summary-2023-2024.html. accessed: 2024-11-13.

Centers for Disease Control and Prevention, 2025. Weekly Hospital Respiratory Data (HRD) Metrics by Jurisdiction, National Healthcare Safety Network (NHSN) (Preliminary). https://data.cdc.gov/Public-Health-Surveillance/Weekly-Hospital-Respiratory-Data-HRD-Metrics-by-Ju/mpgq-jmmr/about_data. Accessed: 2025-01-09.

Chinazzi, M., Davis, J.T., y Piontti, A.P., Mu, K., Gozzi, N., Ajelli, M., Perra, N., Vespignani, A., 2024. A multiscale modeling framework for scenario modeling: Characterizing the heterogeneity of the covid-19 epidemic in the us. Epidemics 47, 100757. doi:10.1016/j.epidem.2024.100757.

Cramer, E., Witthaut, D., Mitsos, A., Dahmen, M., 2023. Multivariate probabilistic forecasting of intraday electricity prices using normalizing flows. Applied Energy 346, 121370. URL: https://www.sciencedirect.com/science/article/pii/S0306261923007341, doi:10.1016/j.apenergy.2023.121370.

Cramer, E.Y., Huang, Y., Wang, Y., Ray, E.L., Cornell, M., Bracher, J., Brennen, A., Rivadeneira, A.J.C., Gerding, A., House, K., Jayawardena, D., Kanji, A.H., Khandelwal, A., Le, K., Mody, V., Mody, V., Niemi, J., Stark, A., Shah, A., Wattanchit, N., Zorn, M.W., Reich, N.G., Gneiting, T., Mühlemann, A., Gu, Y., Chen, Y., Chintanippu, K., Jivane, V., Khurana, A., Kumar, A., Lakhani, A., Mehrotra, P., Pasumarty, S., Shrivastav, M., You, J., Bannur, N., Deva, A., Jain, S., Kulkarni, M., Merugu, S., Raval, A., Shingi, S., Tiwari, A., White, J., Adiga, A., Hurt, B., Lewis, B., Marathe, M., Peddireddy, A.S., Porebski, P., Venkatramanan, S., Wang, L., Dahan, M., Fox, S., Gaither, K., Lachmann, M., Meyers, L.A., Scott, J.G., Tec, M., Woody, S., Srivastava, A., Xu, T., Cegan, J.C., Dettwiller, I.D., England, W.P., Farthing, M.W., George, G.E., Hunter, R.H., Lafferty, B., Linkov, I., Mayo, M.L., Parno, M.D., Rowland, M.A., Trump, B.D., Chen, S., Faraone, S.V., Hess, J., Morley, C.P., Salekin, A., Wang, D., Zhang-James, Y., Baer, T.M., Corsetti, S.M., Eisenberg, M.C., Falb, K., Huang, Y., Martin, E.T., McCauley, E., Myers, R.L., Schwarz, T., Gibson, G.C., Sheldon, D., Gao, L., Ma, Y., Wu, D., Yu, R., Jin, X., Wang, Y.X., Yan, X., Chen, Y., Guo, L., Zhao, Y., Chen, J., Gu, Q., Wang, L., Xu, P., Zhang, W., Zou, D., Chattopadhyay, I., Huang, Y., Lu, G., Pfeiffer, R., Sumner, T., Wang, D., Wang, L., Zhang, S., Zou, Z., Biegel, H., Lega, J., Hussain, F., Khan, Z., Van Bussel, F., McConnell, S., Guertin, S.L., Hulme-Lowe, C., Nagraj, V.P., Turner, S.D., Bejar, B., Choirat, C., Flahault, A., Krymova, E., Lee, G., Manetti, E., Namigai, K., Obozinski, G., Sun, T., Thanou, D., Ban, X., Shi, Y., Walraven, R., Hong, Q.J., van de Walle, A., Ben-Nun, M., Riley, S., Riley, P., Turtle, J., Cao, D., Galasso, J., Cho, J.H., Jo, A., DesRoches, D., Forli, P., Hamory, B., Koyluoglu, U., Kyriakides, C., Leis, H., Milliken, J., Moloney, M., Morgan, J., Nirgudkar, N., Ozcan, G., Piwonka, N., Ravi, M., Schrader, C., Shakhnovich, E., Siegel, D., Spatz, R., Stiefeling, C., Wilkinson, B., Wong, A., Cavany, S., España, G., Moore, S., Oidtman, R., Perkins, A., Ivy, J.S., Mayorga, M.E., Mele, J., Rosenstrom, E.T., Swann, J.L., Kraus, A., Kraus, D., Bian, J., Cao, W., Gao, Z., Ferres, J.L., Li, C., Liu, T.Y., Xie, X., Zhang, S., Zheng, S., Chinazzi, M., Vespignani, A., Xiong, X., Davis, J.T., Mu, K., Piontti, A.P.y., Baek, J., Farias, V., Georgescu, A., Levi, R., Sinha, D., Wilde, J., Zheng, A., Lami, O.S., Bennouna, A., Ndong, D.N., Perakis, G., Singhvi, D., Spantidakis, I., Thayaparan, L., Tsiourvas, A., Weisberg, S., Jadbabaie, A., Sarker, A., Shah, D., Celi, L.A., Penna, N.D., Sundar, S., Berlin, A., Gandhi, P.D., McAndrew, T., Piriya, M., Chen, Y., Hlavacek, W., Lin, Y.T., Mallela, A., Miller, E., Neumann, J., Posner, R., Wolfinger, R., Castro, L., Fairchild, G., Michaud, I., Osthus, D., Wolffram, D., Karlen, D., Panaggio, M.J., Kinsey, M., Mullany, L.C., Rainwater-Lovett, K., Shin, L., Tallaksen, K., Wilson, S., Brenner, M., Coram, M., Edwards, J.K., Joshi, K., Klein, E., Hulse, J.D., Grantz, K.H., Hill, A.L., Kaminsky, K., Kaminsky, J., Keegan, L.T., Lauer, S.A., Lee, E.C., Lemaitre, J.C., Lessler, J., Meredith, H.R., Perez-Saez, J., Shah, S., Smith, C.P., Truelove, S.A., Wills, J., Gardner, L., Marshall, M., Nixon, K., Burant, J.C., Budzinski, J., Chiang, W.H., Mohler, G., Gao, J., Glass, L., Qian, C., Romberg, J., Sharma, R., Spaeder, J., Sun, J., Xiao, C., Gao, L., Gu, Z., Kim, M., Li, X., Wang, Y., Wang, G., Wang, L., Yu, S., Jain, C., Bhatia, S., Nouvellet, P., Barber, R., Gaikedu, E., Hay, S., Lim, S., Murray, C., Pigott, D., Reiner, R.C., Baccam, P., US COVID-19 Forecast Hub Consortium, 2022a. The united states COVID-19 forecast hub dataset. Scientific Data 9, 462. doi:10.1038/s41597-022-01517-w.

Cramer, E.Y., et al., 2022b. Evaluation of individual and ensemble probabilistic forecasts of covid-19 mortality in the united states. Proc. Natl. Acad. Sci. U.S.A. 119, e2113561119. doi:10.1073/pnas.2113561119.

Engebretsen, S., Diz-Lois Palomares, A., Rø, G., Kristoffersen, A.B., Lindstrøm, J.C., Engø-Monsen, K., Kamineni, M., Hin Chan, L.Y., Dale Midtbø, J.E., Stenerud, K.L., Di Ruscio, F., White, R., Frigessi, A., de Blasio, B.F., 2023. A real-time regional model for covid-19: Probabilistic situational awareness and forecasting. PLOS Computational Biology 19, 1–26. URL: https://doi.org/10.1371/journal.pcbi.1010860, doi:10.1371/journal.pcbi.1010860.

FluSight Forecast Hub, 2025. Target Data. https://github.com/cdcepi/FluSight-forecast-hub/tree/main/target-data. Accessed: 2025-01-09.

Gneiting, T., Raftery, A.E., 2007. Strictly proper scoring rules, prediction, and estimation. Journal of the American Statistical Association 102, 359–378. URL: https://doi.org/10.1198/016214506000001437, doi:10.1198/016214506000001437.

Gneiting, T., Stanberry, L.I., Grimit, E.P., Held, L., Johnson, N.A., 2008. Assessing probabilistic forecasts of multivariate quantities, with an application to ensemble predictions of surface winds. TEST 17, 211–235. URL: https://doi.org/10.1007/s11749-008-0114-x, doi:10.1007/s11749-008-0114-x.

Grothe, O., Kächele, F., Krüger, F., 2023. From point forecasts to multivariate probabilistic forecasts: The schaake shuffle for day-ahead electricity price forecasting. Energy Economics 120, 106602. URL: https://www.sciencedirect.com/science/article/pii/S0140988323001007, doi:10.1016/j.eneco.2023.106602.

Held, L., Meyer, S., Bracher, J., 2017. Probabilistic forecasting in infectious disease epidemiology: the 13th armitage lecture. Statistics in Medicine 36, 3443–3460. URL: https://onlinelibrary.wiley.com/doi/abs/10.1002/sim.7363, doi:10.1002/sim.7363.

Howerton, E., Contamin, L., Mullany, L.C., Qin, M., Reich, N.G., Bents, S., Borchering, R.K., Jung, S.m., Loo, S.L., Smith, C.P., Levander, J., Kerr, J., Espino, J., van Panhuis, W.G., Hochheiser, H., Galanti, M., Yamana, T., Pei, S., Shaman, J., Rainwater-Lovett, K., Kinsey, M., Tallaksen, K., Wilson, S., Shin, L., Lemaitre, J.C., Kaminsky, J., Hulse, J.D., Lee, E.C., McKee, C.D., Hill, A., Karlen, D., Chinazzi, M., Davis, J.T., Mu, K., Xiong, X., Pastore y Piontti, A., Vespignani, A., Rosenstrom, E.T., Ivy, J.S., Mayorga, M.E., Swann, J.L., España, G., Cavany, S., Moore, S., Perkins, A., Hladish, T., Pillai, A., Ben Toh, K., Longini, I., Chen, S., Paul, R., Janies, D., Thill, J.C., Bouchnita, A., Bi, K., Lachmann, M., Fox, S.J., Meyers, L.A., Srivastava, A., Porebski, P., Venkatramanan, S., Adiga, A., Lewis, B., Klahn, B., Outten, J., Hurt, B., Chen, J., Mortveit, H., Wilson, A., Marathe, M., Hoops, S., Bhattacharya, P., Machi, D., Cadwell, B.L., Healy, J.M., Slayton, R.B., Johansson, M.A., Biggerstaff, M., Truelove, S., Runge, M.C., Shea, K., Viboud, C., Lessler, J., 2023. Evaluation of the US COVID-19 scenario modeling hub for informing pandemic response under uncertainty. Nature Communications 14, 7260. URL: https://doi.org/10.1038/s41467-023-42680-x, doi:10.1038/s41467-023-42680-x.

Jordan, A., Krüger, F., Lerch, S., 2019. Evaluating probabilistic forecasts with scoringRules. Journal of Statistical Software 90, 1–37. doi:10.18637/jss.v090.i12.

Juul, J.L., Græsbøll, K., Christiansen, L.E., Lehmann, S., 2021. Fixed-time descriptive statistics underestimate extremes of epidemic curve ensembles. Nature Physics 17, 5–8. URL: https://www.nature.com/articles/s41567-020-01121-y, doi:10.1038/s41567-020-01121-y. publisher: Nature Publishing Group.

Kapoor, A., Negi, A., Marshall, L., Chandra, R., 2023. Cyclone trajectory and intensity prediction with uncertainty quantification using variational recurrent neural networks. Environmental Modelling & Software 162, 105654. URL: https://www.sciencedirect.com/science/article/pii/S1364815223000403, doi:10.1016/j.envsoft.2023.105654.

Kendall, M.G., 1938. A New Measure of Rank Correlation. Biometrika 30, 81–93. URL: https://doi.org/10.1093/biomet/30.1-2.81, doi:10.1093/biomet/30.1-2.81.

Loo, S.L., Howerton, E., Contamin, L., Smith, C.P., Borchering, R.K., Mullany, L.C., Bents, S., Carcelen, E., mok Jung, S., Bogich, T., van Panhuis, W.G., Kerr, J., Espino, J., Yan, K., Hochheiser, H., Runge, M.C., Shea, K., Lessler, J., Viboud, C., Truelove, S., 2024. The us covid-19 and influenza scenario modeling hubs: Delivering long-term projections to guide policy. Epidemics 46, 100738. doi:10.1016/j.epidem.2023.100738.

Mathis, S.M., Webber, A.E., León, T.M., Murray, E.L., Sun, M., White, L.A., Brooks, L.C., Green, A., Hu, A.J., Rosenfeld, R., Shemetov, D., Tibshirani, R.J., McDonald, D.J., Kandula, S., Pei, S., Yaari, R., Yamana, T.K., Shaman, J., Agarwal, P., Balusu, S., Gururajan, G., Kamarthi, H., Prakash, B.A., Raman, R., Zhao, Z., Rodríguez, A., Meiyappan, A., Omar, S., Baccam, P., Gurung, H.L., Suchoski, B.T., Stage, S.A., Ajelli, M., Kummer, A.G., Litvinova, M., Ventura, P.C., Wadsworth, S., Niemi, J., Carcelen, E., Hill, A.L., Loo, S.L., McKee, C.D., Sato, K., Smith, C., Truelove, S., Jung, S.m., Lemaitre, J.C., Lessler, J., McAndrew, T., Ye, W., Bosse, N., Hlavacek, W.S., Lin, Y.T., Mallela, A., Gibson, G.C., Chen, Y., Lamm, S.M., Lee, J., Posner, R.G., Perofsky, A.C., Viboud, C., Clemente, L., Lu, F., Meyer, A.G., Santillana, M., Chinazzi, M., Davis, J.T., Mu, K., Pastore y Piontti, A., Vespignani, A., Xiong, X., Ben-Nun, M., Riley, P., Turtle, J., Hulme-Lowe, C., Jessa, S., Nagraj, V.P., Turner, S.D., Williams, D., Basu, A., Drake, J.M., Fox, S.J., Suez, E., Cojocaru, M.G., Thommes, E.W., Cramer, E.Y., Gerding, A., Stark, A., Ray, E.L., Reich, N.G., Shandross, L., Wattanachit, N., Wang, Y., Zorn, M.W., Aawar, M.A., Srivastava, A., Meyers, L.A., Adiga, A., Hurt, B., Kaur, G., Lewis, B.L., Marathe, M., Venkatramanan, S., Butler, P., Farabow, A., Ramakrishnan, N., Muralidhar, N., Reed, C., Biggerstaff, M., Borchering, R.K., 2024. Evaluation of FluSight influenza forecasting in the 2021–22 and 2022–23 seasons with a new target laboratory-confirmed influenza hospitalizations. Nature Communications 15, 6289. URL: https://www.nature.com/articles/s41467-024-50601-9, doi:10.1038/s41467-024-50601-9.

McCabe, R., Kont, M.D., Schmit, N., Whittaker, C., Løchen, A., Walker, P.G.T., Ghani, A.C., Ferguson, N.M., White, P.J., Donnelly, C.A., Watson, O.J., 2021. Communicating uncertainty in epidemic models. Epidemics 37, 100520. URL: https://www.sciencedirect.com/science/article/pii/S1755436521000669, doi:10.1016/j.epidem.2021.100520.

McGowan, C.J., et al., 2019. Collaborative efforts to forecast seasonal influenza in the United States, 2015– 2016. Sci. Rep. 9, 683. URL: https://doi.org/10.1038/s41598-018-36361-9, doi:10.1038/s41598-018-36361-9.

Pic, R., Dombry, C., Naveau, P., Taillardat, M., 2025. Proper scoring rules for multivariate probabilistic forecasts based on aggregation and transformation. Advances in Statistical Climatology, Meteorology and Oceanography 11, 23–58. URL: https://ascmo.copernicus.org/articles/11/23/2025/, doi:10.5194/ascmo-11-23-2025.

Pinson, P., Girard, R., 2012. Evaluating the quality of scenarios of short-term wind power generation. Applied Energy 96, 12–20. URL: https://www.sciencedirect.com/science/article/pii/S0306261911006994, doi:10.1016/j.apenergy.2011.11.004.smartGrids.

Pinson, P., Tastu, J., 2013. Discrimination ability of the Energy score. Number 15 in DTU Compute Technical Report-2013, Technical University of Denmark. URL: https://orbit.dtu.dk/en/publications/discrimination-ability-of-the-energy-score.

Ray, E.L., Brooks, L.C., Bien, J., Biggerstaff, M., Bosse, N.I., Bracher, J., Cramer, E.Y., Funk, S., Gerding, A., Johansson, M.A., Rumack, A., Wang, Y., Zorn, M., Tibshirani, R.J., Reich, N.G., 2023. Comparing trained and untrained probabilistic ensemble forecasts of covid-19 cases and deaths in the united states. International Journal of Forecasting 39, 1366–1383. URL: https://www.sciencedirect.com/science/article/pii/S0169207022000966, doi:10.1016/j.ijforecast.2022.06.005.

Reich, N.G., et al., 2022. Collaborative hubs: Making the most of predictive epidemic modeling. Am. J. Public Health 112, 839–842. URL: https://doi.org/10.2105/AJPH.2022.306831, doi:10.2105/AJPH.2022.306831.

Runge, M.C., Shea, K., Howerton, E., Yan, K., Hochheiser, H., Rosenstrom, E., Probert, W.J., Borchering, R., Marathe, M.V., Lewis, B., Venkatramanan, S., Truelove, S., Lessler, J., Viboud, C., 2024. Scenario design for infectious disease projections: Integrating concepts from decision analysis and experimental design. Epidemics 47, 100775. doi:10.1016/j.epidem.2024.100775.

Scenario Modeling Hub, 2024a. Flu Scenario Modeling Hub. https://fluscenariomodelinghub.org/. Accessed: 2024-02-01.

Scenario Modeling Hub, 2024b. Scenario Modeling Hub. https://scenariomodelinghub.org/. Accessed: 2024-10-08.

Sherratt, K., Srivastava, A., Ainslie, K., Singh, D.E., Cublier, A., Marinescu, M.C., Carretero, J., Garcia, A.C., Franco, N., Willem, L., Abrams, S., Faes, C., Beutels, P., Hens, N., Müller, S., Charlton, B., Ewert, R., Paltra, S., Rakow, C., Rehmann, J., Conrad, T., Schütte, C., Nagel, K., Abbott, S., Grah, R., Niehus, R., Prasse, B., Sandmann, F., Funk, S., 2024. Characterising information gains and losses when collecting multiple epidemic model outputs. Epidemics 47, 100765. URL: https://www.sciencedirect.com/science/article/pii/S1755436524000264, doi:10.1016/j.epidem.2024.100765.

Staid, A., Watson, J.P., Wets, R.J.B., Woodruff, D.L., 2017. Generating short-term probabilistic wind power scenarios via nonparametric forecast error density estimators. Wind Energy 20, 1911–1925. URL: https://onlinelibrary.wiley.com/doi/abs/10.1002/we.2129, doi:10.1002/we.2129.

Székely, G.J., Rizzo, M.L., 2017. The energy of data. Annual Review of Statistics and Its Application 4, 447–479. URL: https://doi.org/10.1146/annurev-statistics-060116-054026, doi:10.1146/annurev-statistics-060116-054026.

Székely, G.J., Rizzo, M.L., 2013. Energy statistics: A class of statistics based on distances. Journal of Statistical Planning and Inference 143, 1249–1272. URL: https://www.sciencedirect.com/science/article/pii/S0378375813000633, doi:10.1016/j.jspi.2013.03.018.

van der Meer, D., 2021. A benchmark for multivariate probabilistic solar irradiance forecasts. Solar Energy 225, 286–296. URL: https://www.sciencedirect.com/science/article/pii/S0038092x2100579X, doi:10.1016/j.solener.2021.07.010.

Wilke, C.O., Bergstrom, C.T., 2020. Predicting an epidemic trajectory is difficult. Proceedings of the National Academy of Sciences of the United States of America 117, 28549–28551. doi:10.1073/pnas.2020200117.

Zhang, Y., Wang, J., Wang, X., 2014. Review on probabilistic forecasting of wind power generation. Renewable and Sustainable Energy Reviews 32, 255–270. URL: https://www.sciencedirect.com/science/article/pii/S1364032114000446, doi:10.1016/j.rser.2014.01.033.

