## Supplementary Material for "Evaluation of stochastic trajectory-based epidemic models using the energy score"

#### Contents

1 2023-24 Flu Scenario Modeling Hub Round 1
2 Other Scores for Model Evaluation 2
3 Weighted Interval Score (WIS) Analysis 3
4 Naive baseline model 6
5 Energy Score Ratio Descriptive Statistics 8
6 Multi-Dimensional Energy Score 10
7 Scenario Modeling Hub Ensemble Model Comparison 11
8 Sampling trajectories in energy score calculation 12
9 Proofs of properness 13

#### 1 2023-24 Flu Scenario Modeling Hub Round

In this paper, we use scenario projections generated by modeling teams during the 2023-24 Flu Scenario Modeling Hub (SMH) projection round. For these projections, modeling teams aimed to predict the behavior of the influenza season under 6 potential future outcomes, with assumptions

| | **Season dominated by influenza A/H3N2. 40% VE against medically attended illness and hospitalization, drops in older age groups.** | **Season dominated by influenza A/H1N1. 40% VE against medically attended illness and hospitalization, similar across all age groups.** |
|---|---|---|
| **Higher than usual vaccine coverage (20% higher than in 2021-22 flu season)** | Scenario A | Scenario B |
| **Business as usual vaccine coverage (same as in 2021-22 flu season)** | Scenario C | Scenario D |
| **Lower than usual vaccine coverage (20% lower than in 2021-22 flu season)** | Scenario E | Scenario F |

Table S1: 2023-24 Flu Scenario Modeling Hub round. Description of the scenarios used by each modeling team in the 2023-24 flu SMH projection round. VE describes assumptions surrounding vaccine effectiveness (VE).

surrounding the vaccine coverage and dominant circulating strain during the 2023-24 influenza season. A description of these scenarios is shown in S1, and incident hospitalization projections were made for each U.S. state, the District of Columbia, and nationally from September 3, 2023 to June 1, 2024 [1]. Ten modeling teams provided projections under these assumptions, contributing 100 stochastic trajectories for each scenario and location. These projections are made prior to the onset of the peak 2023-24 influenza season, and are largely based on historical trends, without much calibration data available for current circulation patterns [1].

#### 2 Other Scores for Model Evaluation

There are a range of known proper scores that can evaluate probabilistic predictions. Metrics applicable to probabilistic forecasts provided as quantiles and full predictive distributions have been widely used for assessing the performance of epidemic projections [2, 3].

For example, the weighted interval score (WIS) is a negatively-oriented proper score applied to $(1-\alpha) \times 100\%$ prediction intervals. The WIS can be decomposed into three components: the width of the prediction interval such that wider intervals are penalized, and a cost if observations lie above or below the prediction interval [2]. The WIS assesses interval forecasts by analyzing the sharpness and calibration of the prediction intervals with respect to the observed data [3]. It is computed at each time point with prediction $P$ and observed value $y$ using the expression

$$\mathrm{WIS}_{\alpha_{0:K}}(P, y) = \frac{1}{K + 0.5}\left( w_0|y - m| + \sum_{k=1}^{K} w_k \mathrm{IS}_{\alpha_k}(P, y) \right), \tag{1}$$

where $\mathrm{IS}_\alpha$ corresponds to the interval score of the $(1 - \alpha)\times 100\%$ prediction interval,

$$\mathrm{IS}_\alpha(P, y) = \underbrace{(u_\alpha - l_\alpha)}_{\text{dispersion}} + \underbrace{\frac{2}{\alpha}(l_\alpha - y)\mathbb{1}(y < l_\alpha)}_{\text{overprediction}} + \underbrace{\frac{2}{\alpha}(y - u_\alpha)\mathbb{1}(y > u_\alpha)}_{\text{underprediction}}, \tag{2}$$

where $K$ is the number of prediction intervals used, $m$ is the median of $P$, and $l_\alpha$ $(u_\alpha)$ is the lower bound (upper bound) of the prediction interval. The standard weights are chosen such that $w_k = \frac{\alpha_k}{2}$ and $w_0 = \frac{1}{2}$. In order to evaluate the performance of a full projection time series using the WIS, we take the average of the WIS calculated at each time point.

Alternatively, the continuous ranked probability score (CRPS) evaluates the cumulative distribution function (CDF) of probabilistic predictions [2, 4]. It is defined as

$$\mathrm{CRPS}(F, y) = \int_{-\infty}^{\infty} [F(x) - \mathbb{1}(x \geq y)]^2 dx,$$

where $F$ is the CDF of the projections and $y$ describes the observed values. The CRPS is a negatively-oriented proper score, and can be strictly proper if the predictive distribution has a finite first moment [5]. It is a generalization of the absolute error, which it reduces to if the projection is a point prediction [4].

Moreover, the CRPS can be approximated by the WIS using properties from the quantile score, given certain weights in the WIS expression [2, 6]. The CRPS also draws comparison to the energy score, which is a multivariate generalization of the CRPS [5]. In the univariate case, where a trajectory is of length one, the energy score and CRPS are identical. This highlights the similarities between how commonly used scoring rules evaluate epidemic projections.

### 3 Weighted Interval Score (WIS) Analysis

To compute the WIS of the Scenario Modeling Hub data, we first estimate the quantiles of the projections using the submitted trajectories for each model, scenario, and location. These quantiles are then used to calculate the WIS based on the corresponding prediction intervals. For consistency, we use the same prediction intervals employed in previous SMH projection rounds, which required quantile-based submissions; namely the 98%, 95%, 90%, 80%,..., and 10% prediction intervals based on 23 submitted quantiles [2]. The WIS is calculated at individual time points, so we compute the average of these scores across all weeks to obtain the WIS for an entire time series.

To compare how the energy score and WIS evaluate probabilistic predictions with respect to one another, we examine how the two scores assess a synthetic predictive distribution at one time point. Note that the energy score calculated at a single time point is equivalent to the CRPS. We generate a negative binomial distribution as our predictive distribution with a mean of 60 and variance of 31, and generate 100 point predictions by randomly drawing from this distribution. We scan a range of observed values that span the predictive distribution, and calculate the energy score and WIS for each observed value. The WIS is found by estimating quantiles from the sampled point predictions, where we use the prediction intervals used by previous rounds of the Scenario Modeling Hub (98%, 95%, 90%, 80%,..., 10%). In Fig. S1A, we show each score with the predictive distribution overlaid. We find that the energy score and WIS both find the lowest (optimal) score at the same observed value, corresponding to the points with the highest probability in the predictive distribution. The scores also diverge away from the optimal score at similar rates as the observed value decreases in likelihood. The energy score has slightly higher values for observations that are in the tail of the predictive distribution. However, as the number of quantiles used to calculate the WIS increases, the two scores will converge. This highlights similarities in the energy score and WIS, and shows that at individual time points, we can expect similarities in how the two scores evaluate a probabilistic prediction.

The WIS is calculated by summing terms that describe the dispersion, or width of the prediction intervals, along with penalty terms if the predictions over or underpredict the observed data. In Fig. S1 B, we show the WIS at each week of the influenza season, as well as the contribution of each term of the expression. In this example, the WIS is being largely penalized for underprediction, where the observations lie above the upper quantiles of the prediction. Similar to the energy score, the WIS is larger near the peak due to its dependence on the magnitude of the surveillance signal. We further highlight similarities between the energy score and WIS in S1C, where we show the correlation between the normalized energy score (Eq. 4 of main text) and normalized WIS. We define the normalized WIS in the same manner as the normalized energy score, where we divide the WIS (that has been averaged across the time series) by the magnitude of the surveillance data for the corresponding time period. We find a strong correlation between the normalized scores, but the correlation is not as strong as what we find with the non-normalized scores shown in Fig. 4D of the main text.

In the main text, we show how the energy score evaluates models with respect to one another. This allows us to rank models in order to see which models perform better or worse. While we find that the energy score and WIS are highly correlated, we are also interested in whether the two scores rank models similarly as well. In Fig. S1D, we show a histogram of the Kendall's $\tau$ rank correlation for the ranking of models for each scenario and location in the 2023-24 Flu SMH hospitalization predictions. A rank correlation of 1 means that the energy score and WIS rank models identically. We find that the rankings are very similar, with a mean rank correlation across models and scenarios of 0.87. This further highlights the similarities in how the WIS and energy score evaluate model performance, but shows that there are cases where the model rankings differ.

Fig. S2 shows the correlation between the normalized energy score and normalized WIS separated for each scenario for the 'MOBS_NEU-GLEAM_FLU' model 2023-24 Flu SMH projections. We find that the correlations between scores are stronger for Scenarios A, C, and E, which correspond to the scenarios assuming an Influenza A/H3N2 dominant season. Examining the trajectories of this model for each scenario (Fig. 3 of main text), we find that the A/H3N2 dominant scenarios (A,C,E) exhibit simple behavior, with a single peak occurring around the same time, with uncertainty in the peak magnitude. The A/H1N1 dominant scenarios (B,D,F) have more complex dynamics, with one or more peaks occurring at different times. This relates to the discussion in the main text related to how the energy score and WIS behave in situations with multi-peak dynamics (Fig. 2 of main text). We see that in the scenarios with simpler dynamics, the correlation between scores is higher than scenarios with variability in the peak timing. This is likely due to the same phenomenon we discuss in the main text, where the energy score and WIS assess complex multi-peak dynamics differently.

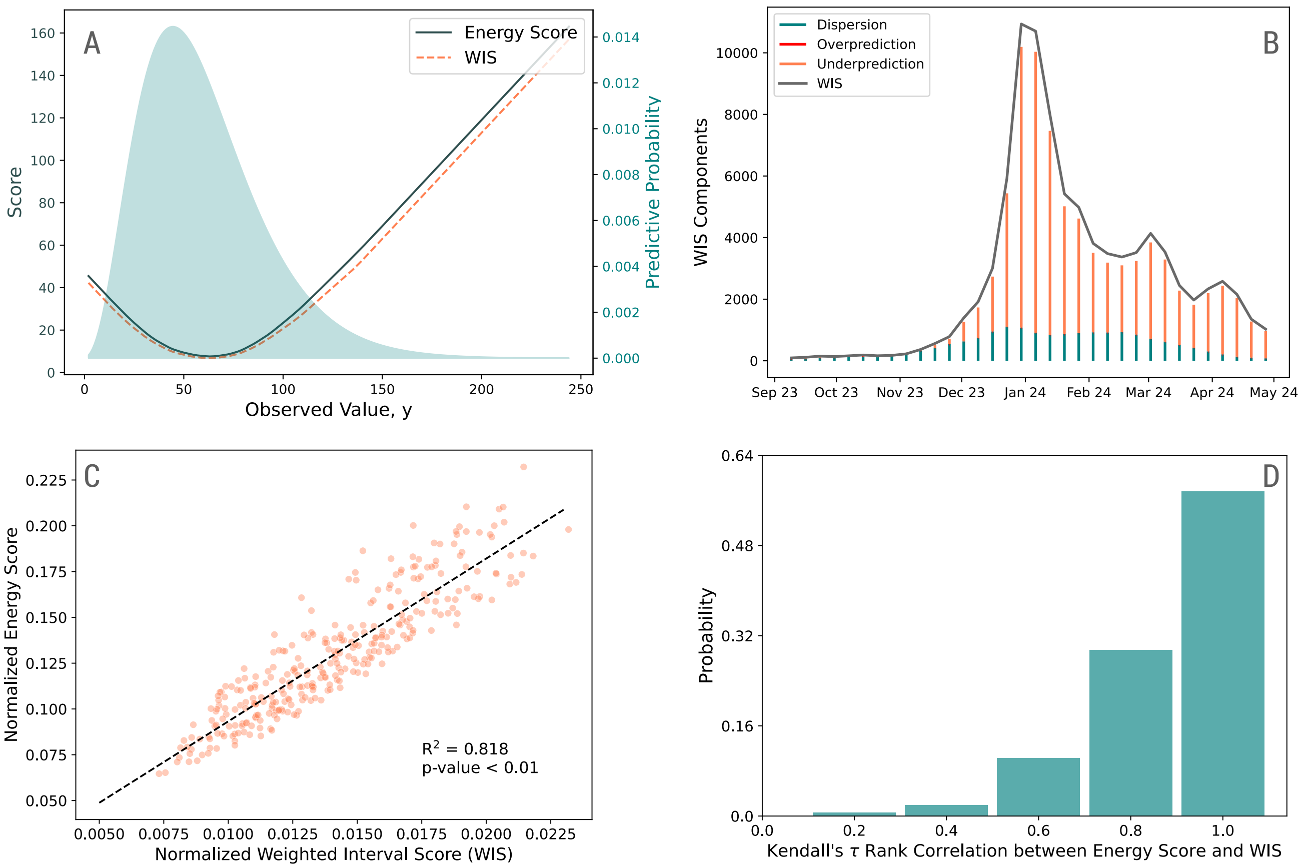


Figure S1: (A) Decomposition of the WIS terms at each week for the 'MOBS_NEU-GLEAM_FLU' model 2023-24 Flu SMH incident hospitalization projections for Scenario D, where blue show the contribution of the dispersion term, red describes the penalty for overprediction, and orange shows the penalty for underprediction. The gray line shows the full score calculated at each week. (B) Comparison of the energy score (gray) and WIS (dashed orange) calculated at one time point as a function of an observed value y given an underlying predictive distribution (blue) with 100 samples drawn from the predictive distribution to calculate the scores. The predictive distribution is a negative binomial distribution with a mean of 60 and variance of 31. (C) Relationship between the normalized WIS and normalized energy score for the 'MOBS_NEU-GLEAM_FLU' model 2023-24 Flu SMH projections. Each point describes the normalized scores for a location and scenario. The WIS was found by estimating the quantiles estimated from the trajectories. Both scores are normalized by dividing the score by the sum of the surveillance data. Dashed line shows the linear regression describing the relationship between the scores. (D) Histogram of the Kendall's $\tau$ rank correlation between the model rankings for the energy score and WIS for each scenario and location for the 2023-24 Flu SMH incident hospitalization projections. A Kendall's $\tau$ rank correlation of 1 means that the energy score and WIS rank models identically.

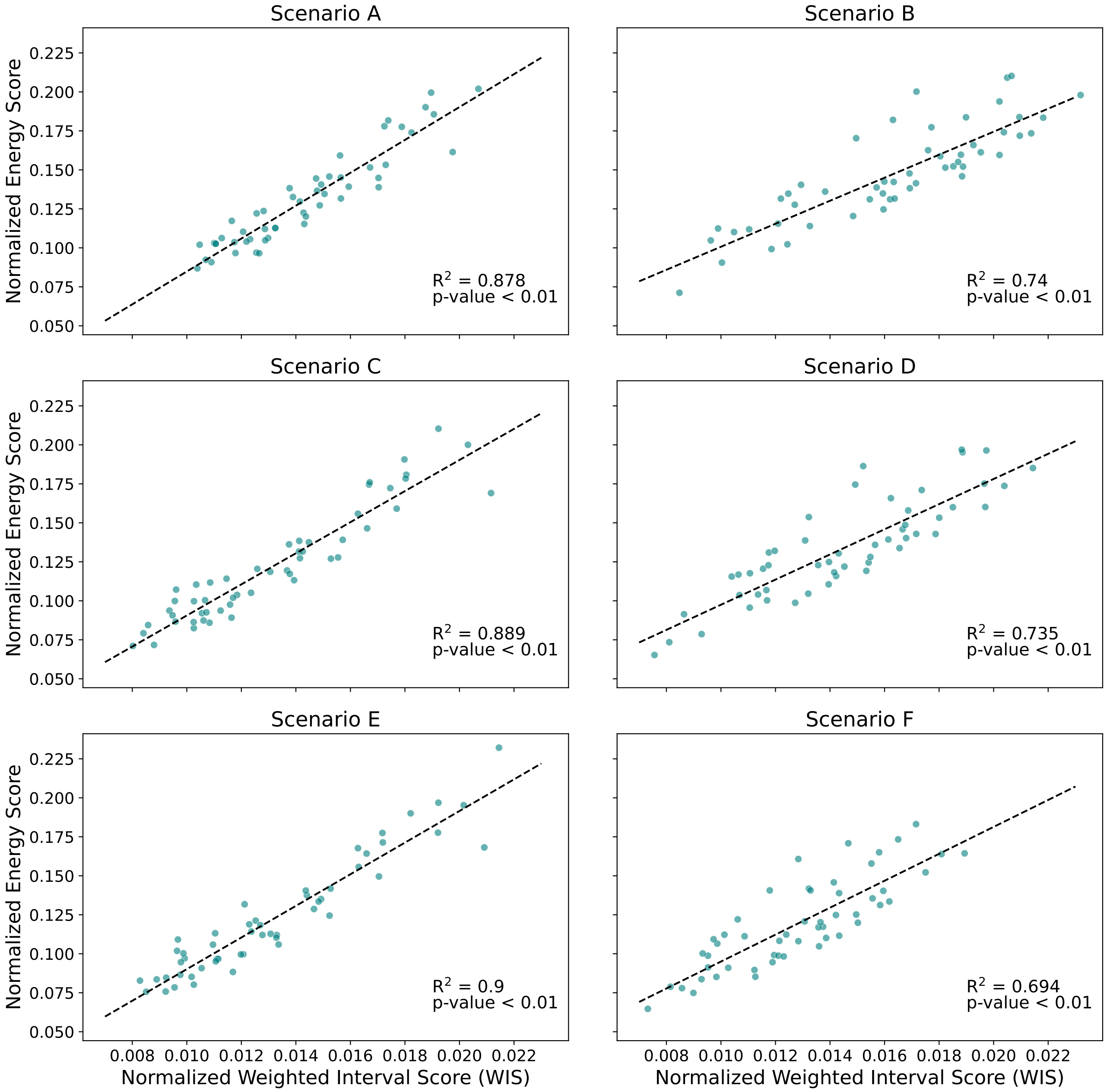


Figure S2: Correlation between the normalized energy score and normalized WIS for each scenario for the 'MOBS_NEU-GLEAM_FLU' model 2023-24 Flu SMH incident hospitalization projections. Each point represents the corresponding scores for a single location for a given scenario. Both scores are normalized by dividing the score by the sum of the surveillance data.

### 4 Naive baseline model

While scoring metrics provide a value against which we can compare models within the Scenario Modeling Hub, this does not actually tell us how well the scenario projections match reality. For example, we can say that a projection with an energy score of 10 performs better than a projection

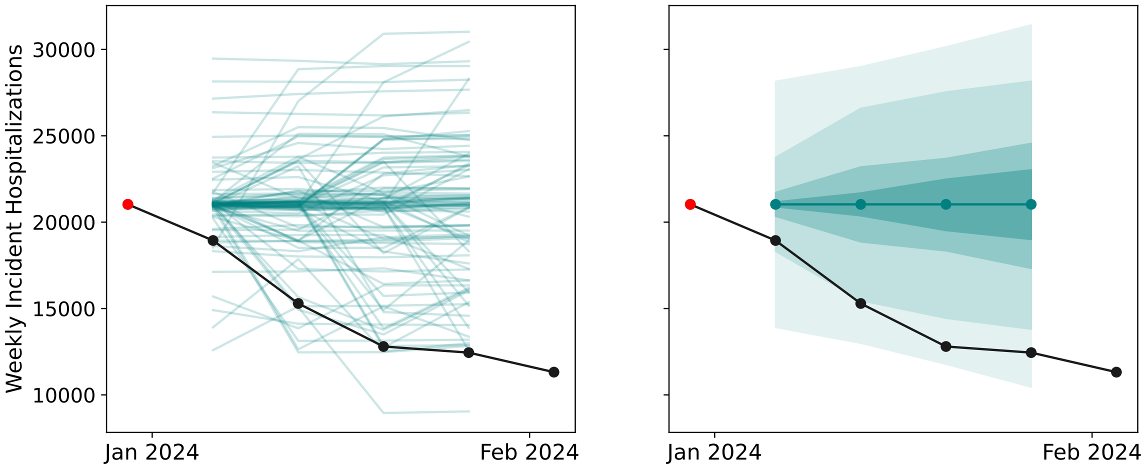


Figure S3: Example of the baseline forecast model for 1-4 week ahead projections given a most recently observed data point of December 30, 2024 (red). Trajectories are generated from the baseline model (left) and can be transformed into quantiles (right). Black line shows observed surveillance data.

with an energy score of 20, but we do not know whether the energy score of 10 is actually a 'good' prediction. Therefore, we employ a naive baseline forecasting model as a reference point against which we compare the performance of the Flu SMH scenario projections. We use the baseline model framework described by Ray et al. where the median is equal to the last observed data point, and the uncertainty around the median is described by historical differences in the changes in incidence counts between consecutive weeks [7]. This creates flat projections of the most recent data, with increasing uncertainty for more distant horizons. Code for the naive baseline model is adapted from the CovidEnsembles (`https://github.com/reichlab/covidEnsembles/`) and CovidModels (`https://github.com/reichlab/covidModels/`) packages in R. To generate trajectories for this model, we create the first horizon (ex. 1-week-ahead) by summing samples drawn from the historical distribution of weekly differences with the most recently observed data point. For longer horizons, the weekly differences are sampled again and summed to the previous horizon projected incidence counts, which generates the expanding uncertainty we observe [7]. We repeat this for the number of weeks ahead being predicted, where the difference from one week to the next is randomly sampled from the distribution of historical differences in incidence counts. Fig. S3 shows an example of the baseline model forecast trajectories and corresponding quantile format.

With this framework, we generate trajectories for 4-week-ahead baseline forecasts for the 2023-24 influenza season in the U.S. We use historical data starting in September 2022 to estimate historical changes in incidence across weeks. Using these trajectories, we calculate the energy score for each U.S. state for the 2023-24 flu season with the 4-week-ahead naive baseline model. In order to have baseline forecasts for each date predicted by the SMH projections, the 4-week-ahead forecasts begin August 12, 2023. In the main text, we compare these scores with the scenario projections to assess whether long-term scenario projections perform better or worse than a short-term baseline forecast.

| | No. locations predicted | Minimum | 5% quantile | Median | 95% quantile | Maximum |
|---|---|---|---|---|---|---|
| Scenario A | 52 | 0.87 | 0.91 | 1.16 | 1.49 | 1.59 |
| Scenario B | 52 | 0.73 | 0.84 | 1.29 | 1.87 | 2.07 |
| Scenario C | 52 | 0.76 | 0.79 | 1.06 | 1.41 | 2.16 |
| Scenario D | 52 | 0.66 | 0.78 | 1.20 | 1.76 | 1.89 |
| Scenario E | 52 | 0.80 | 0.81 | 1.06 | 1.45 | 2.42 |
| Scenario F | 52 | 0.68 | 0.74 | 1.05 | 1.55 | 1.91 |

Table S2: Descriptive statistics of the energy score ratio for the 'MOBS_NEU-GLEAM_FLU' model projections. Table showing the number of locations predicted, minimum, 5% quantile, median, 95% quantile and maximum for energy score ratios across locations for influenza hospitalization predictions for the 'MOBS_NEU-GLEAM_FLU' model of the 2023-24 SMH round across locations for each scenario. The energy score ratio is calculated by dividing the energy score of the SMH model at each scenario and location by the energy score of a 4-week-ahead naive baseline model at each location.

### 5 Energy Score Ratio Descriptive Statistics

In Table S2 we show descriptive statistics of the energy score ratio for the 'MOBS_NEU-GLEAM_FLU' model hospitalization projections across all locations for each scenario of the 2023-24 Flu SMH round. This corresponds to the energy score ratio data used in Fig. 4E in the main text. The energy score ratio compares the energy score of the 'MOBS_NEU-GLEAM_FLU' model with a 4-week-ahead naive baseline model at each scenario and location. An energy score ratio less than 1 corresponds to cases where the 'MOBS_NEU-GLEAM_FLU' model performed better than the baseline model.

In Table S3 we show descriptive statistics of the energy score ratio for each scenario and model across all locations for incident hospitalization projections of the 2023-24 Flu SMH round. This corresponds to the data used in Fig. 5 in the main text, which describes the energy score ratio comparing the energy score of each SMH model to the energy score of a 4-week-ahead naive baseline model. We only show data for the models we used in our analysis, excluding models submitted to the SMH that only reported projections for a single location. Note that there are 52 possible locations included in our analysis: the 50 U.S. states, District of Columbia, and US national projections.

In Table S4, we show descriptive statistics of the energy score ratio for the ensemble model generated by bundling the trajectories of each model together with equal weight. This describes the data used in Fig. 6B in the main text. The energy score ratio compares the behavior of this ensemble with the 4-week-ahead naive baseline model. We include trajectories for all 10 individual models that submitted projections to the 2023-24 Flu SMH, including those that only report scenario projections for a single location.

| Label | Model | No. locations predicted | Minimum | 5% quantile | Median | 95% quantile | Maximum |
|---|---|---|---|---|---|---|---|
| Scenario A | Model I | 52 | 0.87 | 0.91 | 1.16 | 1.49 | 1.59 |
| | Model II | 13 | 0.50 | 0.50 | 0.84 | 1.13 | 1.13 |
| | Model III | 52 | 0.48 | 0.60 | 1.55 | 3.40 | 7.04 |
| | Model IV | 52 | 0.51 | 0.65 | 1.80 | 4.90 | 7.48 |
| | Model V | 52 | 0.93 | 1.09 | 1.79 | 2.52 | 4.70 |
| | Model VI | 51 | 0.66 | 0.87 | 3.19 | 12.22 | 16.66 |
| Scenario B | Model I | 52 | 0.73 | 0.84 | 1.29 | 1.87 | 2.07 |
| | Model II | 13 | 0.79 | 0.81 | 1.02 | 1.55 | 1.61 |
| | Model III | 52 | 0.51 | 0.56 | 0.94 | 2.59 | 4.49 |
| | Model IV | 52 | 0.39 | 0.51 | 1.71 | 3.78 | 4.73 |
| | Model V | 52 | 0.88 | 1.01 | 1.70 | 2.42 | 4.50 |
| | Model VI | 51 | 0.72 | 0.89 | 3.03 | 11.72 | 16.70 |
| Scenario C | Model I | 52 | 0.76 | 0.79 | 1.06 | 1.41 | 2.16 |
| | Model II | 13 | 0.51 | 0.57 | 1.44 | 4.20 | 5.86 |
| | Model III | 52 | 0.54 | 0.66 | 1.79 | 3.91 | 7.87 |
| | Model IV | 52 | 0.50 | 0.79 | 2.31 | 6.02 | 9.35 |
| | Model V | 52 | 0.93 | 1.09 | 1.80 | 2.52 | 4.71 |
| | Model VI | 51 | 0.68 | 0.89 | 2.93 | 12.19 | 16.52 |
| Scenario D | Model I | 52 | 0.66 | 0.78 | 1.20 | 1.76 | 1.89 |
| | Model II | 13 | 0.48 | 0.51 | 0.76 | 2.74 | 3.40 |
| | Model III | 52 | 0.44 | 0.52 | 1.03 | 2.93 | 5.16 |
| | Model IV | 52 | 0.28 | 0.58 | 2.01 | 4.51 | 5.77 |
| | Model V | 52 | 0.88 | 1.01 | 1.70 | 2.42 | 4.50 |
| | Model VI | 51 | 0.65 | 0.82 | 3.07 | 11.84 | 16.58 |
| Scenario E | Model I | 52 | 0.80 | 0.81 | 1.06 | 1.45 | 2.42 |
| | Model II | 13 | 0.45 | 0.71 | 2.75 | 7.51 | 10.00 |
| | Model III | 52 | 0.50 | 0.72 | 2.03 | 4.48 | 8.75 |
| | Model IV | 52 | 0.59 | 1.17 | 2.95 | 7.37 | 11.42 |
| | Model V | 52 | 0.93 | 1.09 | 1.79 | 2.52 | 4.69 |
| | Model VI | 51 | 0.69 | 0.81 | 3.09 | 11.75 | 16.27 |
| Scenario F | Model I | 52 | 0.68 | 0.74 | 1.05 | 1.55 | 1.91 |
| | Model II | 13 | 0.45 | 0.81 | 1.94 | 5.35 | 6.23 |
| | Model III | 52 | 0.51 | 0.54 | 1.22 | 3.36 | 6.09 |
| | Model IV | 52 | 0.26 | 0.86 | 2.35 | 5.46 | 6.74 |
| | Model V | 52 | 0.88 | 1.01 | 1.70 | 2.42 | 4.50 |
| | Model VI | 51 | 0.71 | 0.84 | 3.07 | 11.95 | 16.91 |

Table S3: Descriptive statistics of the energy score ratio for the all analyzed models and scenarios. Table showing the number of locations predicted, minimum, 5% quantile, median, 95% quantile and maximum for energy score ratios across locations for influenza hospitalization predictions for each scenario and model of the 2023-24 SMH round. The energy score ratio is calculated by dividing the energy score of the SMH model at each scenario and location by the energy score of a 4-week-ahead naive baseline model at each location.

| | No. locations predicted | Minimum | 5% quantile | Median | 95% quantile | Maximum |
|---|---|---|---|---|---|---|
| Scenario A | 52 | 0.57 | 0.63 | 1.09 | 2.23 | 2.34 |
| Scenario B | 52 | 0.55 | 0.66 | 1.04 | 1.72 | 1.97 |
| Scenario C | 52 | 0.59 | 0.65 | 1.17 | 2.42 | 2.85 |
| Scenario D | 52 | 0.55 | 0.59 | 1.02 | 1.93 | 2.01 |
| Scenario E | 52 | 0.61 | 0.65 | 1.26 | 2.74 | 3.42 |
| Scenario F | 52 | 0.52 | 0.58 | 1.08 | 2.13 | 2.33 |

Table S4: Descriptive statistics of the energy score ratio for the ensemble model projections. Table showing the number of locations predicted, minimum, 5% quantile, median, 95% quantile and maximum for energy score ratios across locations for influenza hospitalization predictions for the ensemble of trajectories model across locations for each scenario. The energy score ratio is calculated by dividing the energy score of the ensemble model at each scenario and location by the energy score of a 4-week-ahead naive baseline model at each location.

### 6 Multi-Dimensional Energy Score

To calculate the multi-dimensional energy score, we must group trajectories across the different dimensions to create trajectory matrices, where each matrix has one time-series trajectory for each target outcome dimension. The choice of this grouping changes the resulting energy score value. Therefore, trajectories within each matrix should be paired such that trajectories generated by the same simulations are grouped together. While the SMH data is reported with trajectory identifiers, it cannot be ensured that the trajectories across locations are paired. We perform a sensitivity analysis when calculating the multi-dimensional energy score on this data to study the effect of grouping unpaired trajectories. We run multiple iterations of randomizing how these trajectories are paired into the trajectory matrices. In Table S5, we show statistics describing 50 iterations of randomizing the matrices for calculating the multi-dimensional energy score for 'MOBS_NEU-GLEAM_FLU' model for all locations, shown for each scenario. We see that the standard deviation and range of the energy score values is small, with a standard deviation of 0.0002-0.0004 and a range of 0.001-0.002. This suggests that randomly pairing trajectories does not greatly impact the energy score. Moreover, this means that using paired trajectories for the multi-dimensional energy score may not be required if this information is not available. In this example, we also see that it does not impact the ranking of the scenarios, as none of the scores for each scenario overlap with one another.

In the multi-dimensional energy score expression, we introduce a normalization factor, $\Phi^2$, which ensures that outcome dimensions with larger magnitude do not dominate the multi-dimensional energy score value. However, it can be useful to keep the emphasis on outcome dimensions (i.e. locations) with larger signals such that poorly predicting a larger signal is penalized more than dimensions with smaller signals. We compare results of the multi-dimensional energy score with and without the normalization factor, $\Phi_j^2$, in Fig. S4. To calculate the multi-dimensional energy score with no normalization factor, we simply remove $\Phi_j^2$ from the expression (i.e. $\Phi_j^2$=1) in Eq. 5 of the main text. Similar to Fig. 4E of the main text, we compute the non-normalized multi-

| | Mean | Standard Deviation | Minimum | Maximum | Range |
|---|---|---|---|---|---|
| Scenario A | 0.868 | 0.0003 | 0.867 | 0.868 | 0.002 |
| Scenario B | 1.005 | 0.0002 | 1.004 | 1.005 | 0.001 |
| Scenario C | 0.843 | 0.0004 | 0.841 | 0.843 | 0.002 |
| Scenario D | 0.917 | 0.0002 | 0.917 | 0.918 | 0.001 |
| Scenario E | 0.857 | 0.0004 | 0.856 | 0.858 | 0.002 |
| Scenario F | 0.816 | 0.0003 | 0.815 | 0.817 | 0.001 |

Table S5: Descriptive statistics of the multi-dimensional energy score for the 'MOBS_NEU-GLEAM_FLU' model. Table showing the mean, standard deviation, minimum, maximum, and range for the multi-dimensional energy score across locations for the 'MOBS_NEU-GLEAM_FLU' model influenza hospitalization predictions for each scenario of the 2023-24 SMH round, given 50 iterations of randomly pairing trajectories.

dimensional energy score for 50 iterations of randomly shuffling the trajectory identifiers used to pair trajectories, and create an energy score ratio comparing the 'MOBS_NEU-GLEAM_FLU' model to the 4-week-ahead naive baseline model. The blue star in Fig. S4 represents the median non-normalized multi-dimensional energy score ratio, which can be compared to the red star, which shows the multi-dimensional score with normalization included. We find that the energy score ratios are quite similar, and the deviations from the normalized score are likely a consequence of scores from large states biasing the calculation.

### 7 Scenario Modeling Hub Ensemble Model Comparison

In the main text, we describe a method of generating an ensemble model where we simply bundle all trajectories of component models and assign equal weight to each one. The SMH reports 3 ensemble models with projections of the submitted modeling teams. These ensemble models are generated using the Vincent average and linear opinion pool (LOP) methods, where the LOP method is constructed in both a trimmed, where extreme values are excluded, and untrimmed manner [3]. The 3 resulting ensemble models are called the Ensemble_vincent, Ensemble_LOP, and Ensemble_LOP_untrimmed models. These models are calculated and reported in quantile format. We are interested in comparing the difference between the SMH ensemble models and the ensemble of trajectories model analyzed in the main text. Since the SMH ensemble models are only reported as quantiles (and not trajectories), we use the WIS for this analysis. In Fig. S5, we show the relationship between the WIS of each SMH ensemble model and the WIS of the ensemble of trajectories model. The WIS for the trajectory ensemble is calculated by estimating quantiles from the trajectories before computing the WIS. We find a strong positive correlation between the trajectories ensemble and all three SMH ensemble models. However, the Ensemble_LOP_untrimmed model has the strongest correlation with the ensemble of trajectories method. This is reasonable since both methods account for the full uncertainty of all projections, where the Ensemble_vincent and Ensemble_LOP models reduce this variation. This demonstrates the trajectories ensemble performance in

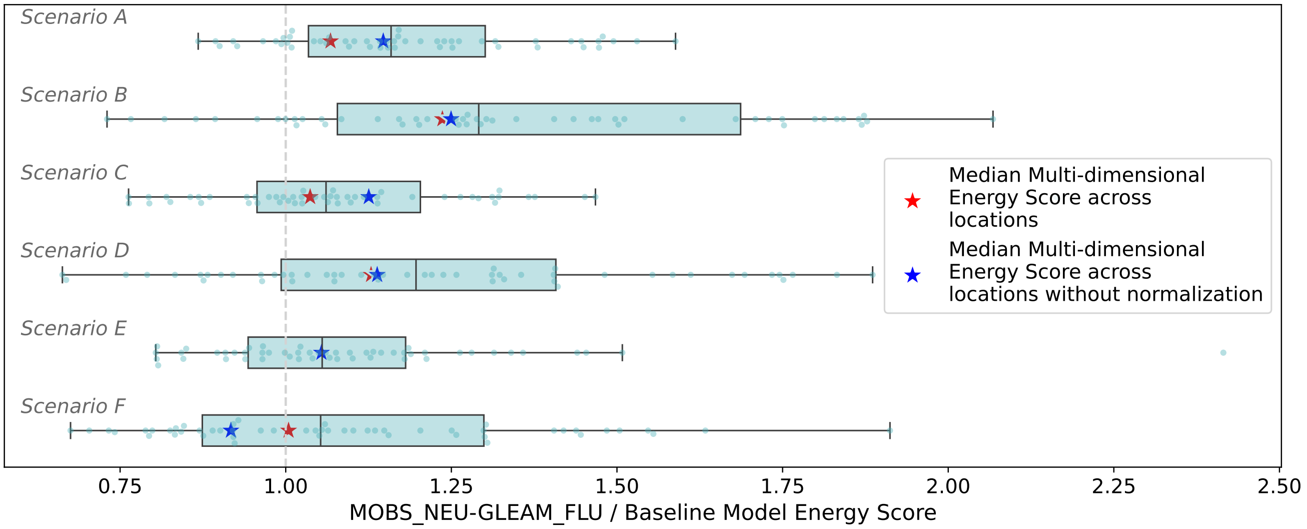


Figure S4: Boxplot of energy score ratio across 52 locations for the 'MOBS_NEU-GLEAM_FLU' model compared to a 4-week-ahead naive baseline model for each scenario. Vertical dashed line shows where the 'MOBS_NEU-GLEAM_FLU' and baseline model have the same scores, where ratios below one describe when the model performs better than the baseline. The overlaid red stars show the median of the multi-dimensional energy score ratio across locations for 50 iterations of randomizing the trajectory pairings. The blue stars show the multi-dimensional energy score without the use of a normalization factor. Boxplots are created such that the box shows the 25%, 50% and 75% quantiles, and the whiskers represent 1.5 × IQR.

comparison to the SMH ensemble models. Choosing different weighting of trajectories within the ensemble of trajectories could change this uncertainty and allow for many other variations of using trajectories for ensemble model generation.

### 8 Sampling trajectories in energy score calculation

Given the pairwise comparison between all pairs of trajectories in order to calculate the second term of the energy score, this computation could become infeasible if we are looking at thousands or millions of trajectories from a model's output. We propose the use of sampling from the full group of trajectories if it is intractable to find the energy score using all of them. We show the outcome of sampling trajectories using the 'MOBS_NEU-GLEAM_FLU' model for the 2023-24 Round 1 of the Flu Scenario Modeling Hub (SMH). To do this, we randomly sample $n$ trajectories for each location and scenario out of the 100 trajectories submitted, and we calculate the energy score using these $n$ trajectories. We repeat this process for 50 iterations and compare the results with the energy score found by using the full set of trajectories.

In Fig. S6 A, we show how the number of sampled trajectories used in the energy score calculation impacts how close our estimated energy score is to the true value. This is shown across 50 iterations of sampling trajectories for four different densities of samples. We find good agreement even for very low number of trajectories sampled, and just 10% of trajectories is sufficient to get a near perfect estimation of the true energy score, with an $R^2 = 0.981$. If we average across these

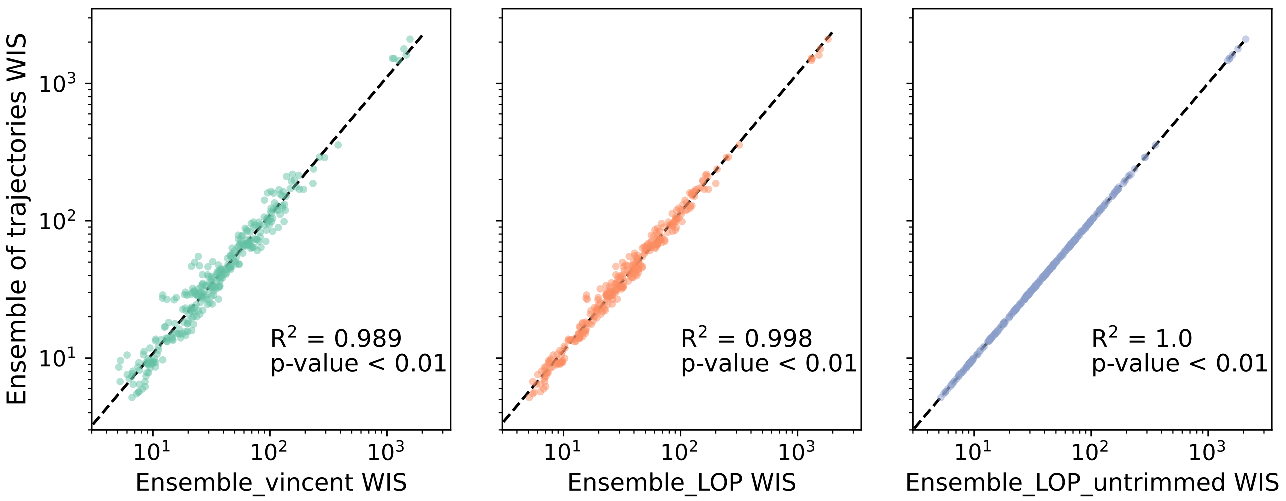


Figure S5: Correlation between the WIS of the ensemble models reported by the SMH (x-axis) and the WIS of the ensemble of trajectories method (y-axis). Each subplot shows one of the 3 different SMH-reported ensemble models with Ensemble_vincent (left), Ensemble_LOP (middle), and Ensemble_LOP_trimmed (right). Each point represents the corresponding WIS scores for a given scenario and location. Dashed black line shows the linear regression fit, with the $R^2$ and two-sided p-value describing the correlation.

iterations, we can improve our estimation for a lower percentage of trajectories sampled. Fig. S6 B shows that with only 5% of trajectories used in the energy score calculation, we get an energy score nearly identical to the truth. This highlights the utility of sampling in calculating the energy score when the total number of stochastic trajectories may be computationally challenging. Even when taking a small sample of trajectories, we can find a close estimate of the true energy score.

### 9 Proofs of properness

In this section we will show proofs that the energy score is a strictly proper scoring rule and that the weighted interval score (WIS) is a proper score. Let us begin with a few important definitions that will be used in both proofs. Let $S(F, y)$ be a score for a probability distribution function $F$ that gives probabilistic predictions of some target $x$. Let $y$ be the observed surveillance data of the prediction target of interest, and $G$ the true underlying distribution for $y$. We call $E_G[S(F, y)] = S(F)$ the expected score. A negatively-oriented score is such that smaller scores correspond to better predictions. Our definitions of the energy score and WIS are both negatively-oriented scores. Given these definitions, we can say that $S$ is a negatively-oriented proper score if $S(G) \leq S(F)$ for all $F$. Moreover, $S$ is a negatively oriented strictly proper score if $S(G) \leq S(F)$ for all $F$, with equality if and only if $F = G$. Therefore, to prove that a score $S$ is proper, we must show that $S(F) - S(G) \geq 0$. To prove that a score is strictly proper, we must show that $S(F) - S(G) \geq 0$ with equality if and only if $F = G$.

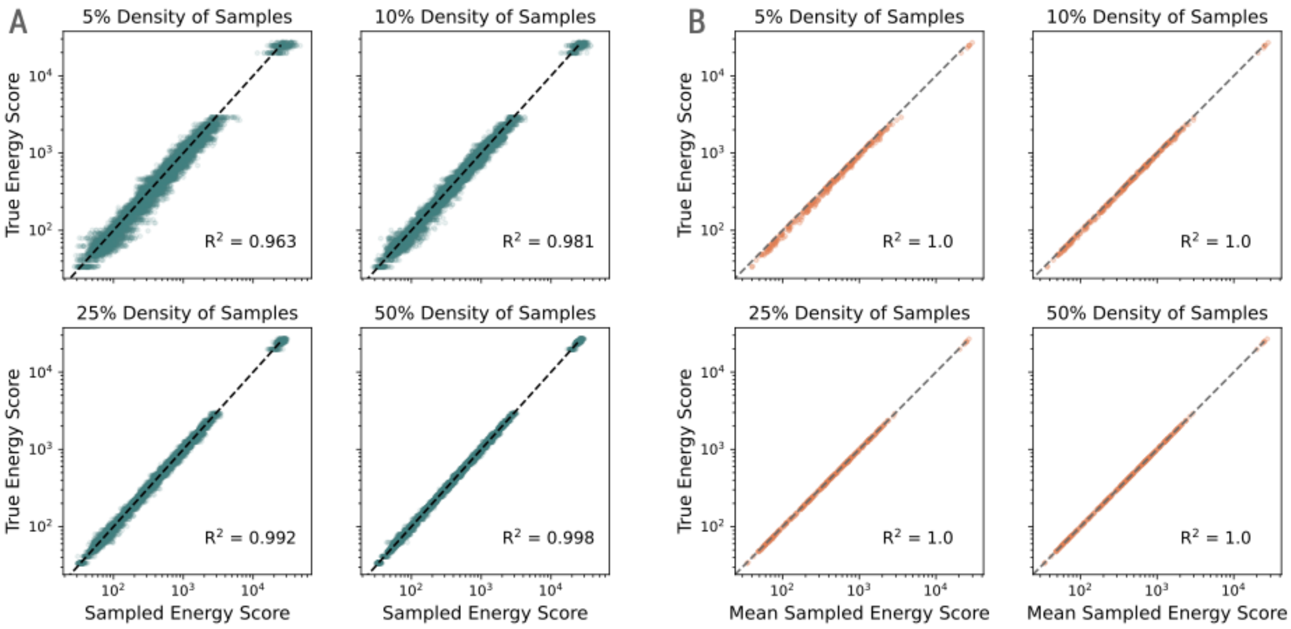


Figure S6: (A) Sampled energy score compared to the true energy score for different densities of sampled trajectories (5, 10, 25, and 50 %) for the 'MOBS_NEU-GLEAM_FLU' model. Dots show a single sampled energy score compared to its corresponding true value for a given location and scenario. (B) Sampled energy score averaged across 50 iterations compared to the true energy score for different densities of sampled trajectories (5, 10, 25, and 50 %) for the 'MOBS_NEU-GLEAM_FLU' model. Dots show the average sampled energy score compared to its corresponding true value for a given location and scenario. Black dotted line shows the y=x line where the sampled and true energy scores are equivalent.

#### Proof that the energy score is strictly proper

To show that the energy score $ES(F, y)$ is strictly proper, we need to show that the expected energy score is uniquely minimized when the prediction distribution $F$ equals the distribution $G$ of the observed value. More formally, we want to show that

$$E_G[\mathrm{ES}(F, y)] \equiv \mathrm{ES}(F) \geq \mathrm{ES}(G) \quad \forall F \tag{3}$$

where the equality holds if and only if $F = G$.

From Refs. [5, 8] we can show that for $y$ a $d$-dimensional vector, the energy score is

$$\mathrm{ES}(F, y) = A \int_{\mathbb{R}^d} \frac{||\phi_F(v) - e^{iv\cdot y}||^2}{||v||^{d+1}} \mathrm{d}v \,, \tag{4}$$

where $\phi_F(v) \equiv E_F\left[e^{iv\cdot x}\right]$ is the characteristic function of $F$, $v$ is also a $d$-dimensional vector, $||\cdot||$ is the Euclidean distance, and

$$A = -\frac{\Gamma\left(\frac{d+1}{2}\right)}{2\pi^{\frac{d+1}{2}}} \,.$$

This shows that the energy score calculates the distance between the characteristic function between the prediction $F$ and the characteristic function of the observed data [5]. Given this, the expected energy score corresponds to:

$$E_G\left[\mathrm{ES}(F,y)\right] = A \cdot E\left[\int_{\mathbb{R}^d} \frac{||\phi_F(v) - e^{iv\cdot y}||^2}{||v||^{d+1}} \mathrm{d}v\right] . \tag{5}$$

To simplify the numerator, we use the fact that the squared norm of a complex number $z$ is given by $||z||^2 = z\bar{z}$, where $\bar{z}$ is the complex conjugate of $z$. If we apply this to our expression of the expected energy score we find

$$= A \cdot E\left[\int_{\mathbb{R}^d} \frac{(\phi_F(v) - e^{iv\cdot y})(\bar{\phi}_F(v) - e^{-iv\cdot y})}{||v||^{d+1}} \mathrm{d}v\right] .$$

If we multiply through all terms and take the expectation over $y$ we get:

$$\begin{aligned}
&= A \cdot E\left[\int_{\mathbb{R}^d} \left(\frac{\phi_F(v)\bar{\phi}_F(v) - e^{iv\cdot y}\bar{\phi}_F(v) - e^{-iv\cdot y}\phi_F(v) + 1}{||v||^{d+1}}\right) \mathrm{d}v\right] , \\
&= A \int_{\mathbb{R}^d} \left(\frac{\phi_F(v)\bar{\phi}_F(v) - \phi_G(v)\bar{\phi}_F(v) - \bar{\phi}_G(v)\phi_F(v) + 1}{||v||^{d+1}}\right) \mathrm{d}v ,
\end{aligned}$$

where $\phi_G(v)$ is the characteristic function of $G$ describing the observed data. If we add and subtract $\phi_G(v)\bar{\phi}_G(v)$ from the numerator inside the integral, we can separate the integral in two parts, to find

$$\begin{aligned}
&= A \int_{\mathbb{R}^d} \left(\frac{\phi_F(v)\bar{\phi}_F(v) - \phi_G(v)\bar{\phi}_F(v) - \bar{\phi}_G(v)\phi_F(v) + \phi_G(v)\bar{\phi}_G(v)}{||v||^{d+1}}\right) \mathrm{d}v \\
&+ A \int_{\mathbb{R}^d} \frac{1 - \phi_G(v)\bar{\phi}_G(v)}{||v||^{d+1}} \mathrm{d}v ,
\end{aligned}$$

If we gather the terms and use the complex conjugate definition of the norm, we can rewrite the numerator as a squared Euclidean distance and show that the expected energy score is

$$E_G\left[\mathrm{ES}(F,y)\right] = A \int_{\mathbb{R}^d} \frac{||\phi_F(v) - \phi_G(v)||^2}{||v||^{d+1}} \mathrm{d}v + B , \tag{6}$$

where $B = A \int_{\mathbb{R}^d} \frac{1-\phi_G(v)\bar{\phi}_G(v)}{||v||^{d+1}} \mathrm{d}v$ is the second integral described above. Since $B$ does not depend on the predictions $F$, we must only look at the first term in order to minimize the expected energy score. This shows that the expected energy score is minimized if and only if $\phi_F(v) = \phi_G(v)$. Since the characteristic functions $\phi_F(v)$ and $\phi_G(v)$ uniquely characterize the underlying probability distributions, this condition is equivalent to $F = G$, which proves the energy score is strictly proper. □

#### Proof that WIS is proper

For a prediction composed of quantiles, we can say that the expected score given the predicted quantiles $r_i$ and probability distribution of the observed data $G$ is

$$E_G[S(r_1, r_2; y)] = S(r_1, ..., r_k; G).$$

We draw comparison to a general quantile scoring rule to show that the WIS is proper [5].

Ref. [5] shows that for predicting a single quantile, the scoring rule

$$S(r; y) = \alpha s(r) + (s(y) - s(r))\mathbb{1}(y \leq r)] + h(y) \tag{7}$$

is proper for quantile levels $\alpha \in (0, 1)$, $s$ is a non-decreasing functions describing the score, $r$ is the predicted quantile, and $h(y)$ is an arbitrary function. First, we will show that a score of this form is proper, using Theorem 6 from Ref. [5] and then we show how the interval score can be written in a general form of this example.

Now we will show that Eq. 7 is proper for $s$ non-decreasing. To do this, we need to show that for a score $S(r; y)$ with predicted quantile $r$, the expected score is maximized by the true quantile $q$ of $G$ at level $\alpha$. In this proof, we show the case for a score $S$ that is positively-oriented, where larger scores are better. This means that the score is proper if it is maximized by the underlying distribution of the observed values. The same proof can be shown for a negatively-oriented score by multiplying all terms by negative one. The expected score can be found by calculating

$$S(r_1, r_2; G) = \int S(r_1, r_2; y)\partial\mu_G(y),$$

where $\mu_G(y)$ this probability measure for the observed data distribution $G$ [5]. Let the probability measure be such that $\mu_G(q) = \alpha$. Therefore, a scoring rule in the form of Eq. 7 is proper if $S(q; G) \geq S(r; G)$. In this example, we assume that $r < q$. The score $S(r; y)$ can be written as

$$S(r; y) = \alpha s(r) + (s(y) - s(r))\mathbb{1}(y \leq r) + h(y). \tag{8}$$

Therefore, the expected score given predicted quantiles $r$ is

$$S(r; G) = \int_0^\infty [\alpha s(r) + (s(y) - s(r))\mathbb{1}(y \leq r) + h(y)]\, \partial\mu_G(y),$$

if we consider $y \in [0, \infty)$. We take the integral of the term multiplied by the indicator function from 0 to $r$ due to the indicator function restriction, simplify, and find

$$S(r; G) = \alpha s(r) - s(r)\mu_G(r) + \int_0^r s(y)\partial\mu_G(y) + \int_0^\infty h(y)\partial\mu_G(y).$$

To show that the true quantile maximizes this score, we subtract this expression from the score calculated using the true quantiles $q$. Using the same methodology as above, the expected score of the true quantiles is

$$S(q; G) = \alpha s(q) - s(q)\mu_G(q) + \int_0^q s(y)\partial\mu_G(y) + \int_0^\infty h(y)\partial\mu_G(y) = \int_0^q s(y)\partial\mu_G(y) + \int_0^\infty h(y)\partial\mu_G(y).$$

The first two terms cancel since $\mu_G(q) = \alpha$. Now, if we subtract the expected score of the true and predicted quantiles, we find

$$S(q;G) - S(r;G) = \int_0^q s(y)\partial\mu_G(y) + \int_0^\infty h(y)\partial\mu_G(y) - \Bigg[\alpha s(r) - s(r)\mu_G(r) + \int_0^r s(y)\partial\mu_G(y) + \int_0^\infty h(y)\partial\mu_G(y)\Bigg],$$

by canceling the $h(y)$ term and rearranging terms, we get

$$S(q;G) - S(r;G) = s(r)\mu_G(r) - \alpha s(r) + \int_0^q s(y)\partial\mu_G(y) - \int_0^r s(y)\partial\mu_G(y).$$

Now we can combine the integrals since $r < q$, and find that

$$S(q;G) - S(r;G) = s(r)\mu_G(r) - \alpha s(r) + \int_r^q s(y)\partial\mu_G(y).$$

Since $s(y)$ is non-decreasing, then $s(y) \geq s(r)$ for $y \in [r, q]$. Therefore, $\int s(y)d\mu_G(y) \geq \int s(r)\partial\mu_G(y) = s(r)(\mu_G(q) - \mu_G(r))$. Using this and the fact that $\mu_G(q) = \alpha$,

$$S(q;G) - S(r;G) \geq s(r)(\mu_G(q) - \mu_G(r)) + s(r)\mu_G(r) - \alpha s(r) = 0.$$

Therefore, $S(q;G) \geq S(r;G)$, meaning that the expected score is maximized by the observed values. Note that this is the case for $r < q$, but a similar proof would follow for the alternative case. This shows that a score of the form Eq. 8 is a proper score, since the expected score is maximized by the true quantiles. To translate this to a case with a negatively-oriented score, we simply must negate the expression above and show that the score is minimized by its true quantiles.

This score can be written in a more general case for predicting $k$ quantiles at levels $\alpha_1, ...\alpha_k \in (0, 1)$ using the expression

$$S(r_1, ..., r_k; y) = \sum_{i=1}^{k} [\alpha_i s_i(r_i) + (s_i(y) - s_i(r_i))\mathbb{1}(y \leq r_i)] + h(y), \tag{9}$$

where it is proper if $s_i$ are non-decreasing [5]. This proof is an extension of the proof shown above for predicting multiple quantiles.

We can write the interval score in the form presented in Ref. 9 where $\alpha_1 = \frac{\alpha}{2}$, $\alpha_2 = 1 - \frac{\alpha}{2}$, $s_1(y) = s_2(y) = 2\frac{y}{\alpha}$, and $h(y) = -2\frac{y}{\alpha}$, as described in [5]. Note that this will give us the negated version of the typical, negatively oriented interval score [2]. Plugging in these functions into Eq. 9, we find that the interval score is

$$IS = r_1 + \frac{2}{\alpha}(y - r_1)\mathbb{1}(y \leq r_1) + 2\frac{r_2}{\alpha} - r_2 + \frac{2}{\alpha}(y - r_2)\mathbb{1}(y \leq r_2) - 2\frac{y}{\alpha}.$$

Rearranging terms and simplifying, we get

$$IS = (r_1 - r_2) + \frac{2}{\alpha}(y - r_1)\mathbb{1}(y \leq r_1) + \frac{2}{\alpha}(r_2 - y)\mathbb{1}(y > r_2).$$

To get the expression for the interval score, we let $r_1 = l$, $r_2 = u$ the lower and upper quantiles of the prediction, respectively. This can be written as

$$IS = (l - u) + \frac{2}{\alpha}(y - l)\mathbb{1}(y \leq l) + \frac{2}{\alpha}(u - y)\mathbb{1}(y > u).$$

This shows that the interval score can be written in the generalized form of Eq. 9, meaning that it is a proper score. This is the positively-oriented formulation of the interval score, but to write the typical, negatively-oriented expression we must multiply all terms by -1. Since the weighted interval score is a linear combination of interval scores, it is also proper because a sum across proper scores is still proper [2].

### References


[1] Scenario Modeling Hub. Flu Scenario Modeling Hub; 2024. `https://fluscenariomodelinghub.org/`.

[2] Bracher J, Ray EL, Gneiting T, Reich NG. Evaluating epidemic forecasts in an interval format. PLOS Computational Biology. 2021;17(2):1–15. doi:10.1371/journal.pcbi.1008618.

[3] Howerton E, Contamin L, Mullany LC, Qin M, Reich NG, Bents S, et al. Evaluation of the US COVID-19 Scenario Modeling Hub for informing pandemic response under uncertainty. Nature Communications. 2023;14(1):7260. doi:10.1038/s41467-023-42680-x.

[4] Gneiting T, Balabdaoui F, Raftery AE. Probabilistic Forecasts, Calibration and Sharpness. Journal of the Royal Statistical Society Series B: Statistical Methodology. 2007;69(2):243–268. doi:10.1111/j.1467-9868.2007.00587.x.

[5] Gneiting T, Raftery AE. Strictly Proper Scoring Rules, Prediction, and Estimation. Journal of the American Statistical Association. 2007;102(477):359–378. doi:10.1198/016214506000001437.

[6] Gneiting T, Ranjan R. Comparing Density Forecasts Using Threshold- and Quantile-Weighted Scoring Rules. Journal of Business & Economic Statistics. 2011;29(3):411–422. doi:10.1198/jbes.2010.08110.

[7] Ray EL, Brooks LC, Bien J, Biggerstaff M, Bosse NI, Bracher J, et al. Comparing trained and untrained probabilistic ensemble forecasts of COVID-19 cases and deaths in the United States. International Journal of Forecasting. 2023;39(3):1366–1383. doi:https://doi.org/10.1016/j.ijforecast.2022.06.005.

[8] Székely GJ, Rizzo ML. Energy statistics: A class of statistics based on distances. Journal of Statistical Planning and Inference. 2013;143(8):1249–1272. doi:https://doi.org/10.1016/j.jspi.2013.03.018.
